# Directed exploration is reduced by an aversive interoceptive state induction in healthy individuals but not in those with affective disorders

**DOI:** 10.1101/2024.06.19.24309110

**Authors:** Ning Li, Claire A. Lavalley, Ko-Ping Chou, Anne E. Chuning, Samuel Taylor, Carter M. Goldman, Taylor Torres, Rowan Hodson, Robert C. Wilson, Jennifer L. Stewart, Sahib S. Khalsa, Martin P. Paulus, Ryan Smith

## Abstract

Elevated anxiety and uncertainty avoidance are known to exacerbate maladaptive choice in individuals with affective disorders. However, the differential roles of state vs. trait anxiety remain unclear, and underlying computational mechanisms have not been thoroughly characterized. In the present study, we investigated how a somatic (interoceptive) state anxiety induction influences learning and decision-making under uncertainty in individuals with clinically significant levels of trait anxiety. A sample of 58 healthy comparisons (HCs) and 61 individuals with affective disorders displaying elevated anxiety symptoms (iADs; i.e., anxiety and/or depression) completed a previously validated explore-exploit decision task, with and without an added breathing resistance manipulation designed to induce state anxiety. Computational modeling revealed a significant group-by-condition interaction, such that information-seeking (i.e., directed exploration) in HCs was reduced by the anxiety induction (Cohen’s *d*=.47, *p*=.013), while no change was observed in iADs. The iADs also showed slower learning rates than HCs across conditions (Cohen’s *d*=.52, *p*=.003), suggesting their uncertainty decreased more slowly over time. These findings highlight a complex interplay between trait anxiety and state anxiety. Specifically, state anxiety may attenuate reflection on uncertainty in healthy individuals, while familiarity with anxious states in those with high trait anxiety may create an insensitivity to this effect.

## Introduction

Persistent uncertainty and maladaptive avoidance are key maintenance factors in anxiety disorders and a major focus of psychotherapy (1, 2). While considerable progress has been made in understanding the neural and cognitive mechanisms associated with anxiety, underlying computational processes have only been examined in a limited number of studies to date (3, 4). For example, subclinical levels of trait anxiety have been associated with reduced flexibility in learning rates when there is a change in the stability of environmental statistics (5), and individuals with anxiety disorders show elevated learning rates in general (6), suggesting the belief that the environment is volatile or that action-outcome contingencies often change unexpectedly. Both depression and anxiety have also been associated with elevated learning from punishment in particular (7). More recently, trait anxiety has been linked to a greater tendency to infer changes in the underlying causes of aversive outcomes during extinction learning – which may facilitate return of fear and reduce the long-term efficacy of behavioral therapies (8).

While such studies provide important insights into the learning processes that may contribute to depression and anxiety disorders, they do not fully account for avoidance or other behaviors driven by intolerance of uncertainty. There could also be multiple computational mechanisms underlying avoidance behavior, each representing distinct hypotheses and possible treatment targets. For example, individuals must often seek information to learn that a feared situation is tolerable. Avoidance prevents such learning, which can in turn maintain both anxiety and further avoidance. This speaks to the explore-exploit dilemma (9, 10), which has begun to receive attention in psychiatry and substance use research (11–13). This dilemma reflects the need to judge whether one has sufficient information to maximize reward (or minimize punishment), or whether one should first seek more information. In avoidance, an individual may hold the confident belief that feared situations are dangerous, while, in fact, exploration would allow them to learn otherwise. Yet, there are multiple types of exploration, and factors that could deter exploration, which have not been thoroughly evaluated.

To date, only a few studies have examined the relationship between affective disorders and information-seeking. For example, one study found that ‘directed’ exploration (DE) in a community sample was lower in those with higher negative affective symptoms (14), while another study associated lower DE with greater somatic anxiety in particular (15). This type of exploration is strategically directed toward situations and actions for which an individual has had fewer past experiences. Further studies examining effects of traumatic/unpredictable childhood environments (16, 17) – which often correlate with affective disorders (18–20) – also suggest negative effects on DE. Other supportive work has shown that: higher stress/anxiety is associated with less exploration in a virtual-reality plus-maze (21), agoraphobia and anxiety sensitivity are associated with less exploratory behavior (22), and increases in cortisol in response to an acute stressor and scores on a chronic stress questionnaire are both associated with under-exploration in foraging tasks (23). Aversive arousal states are also generally known to reduce cognitive control network activity in neuroscience studies (24–28), consistent with reduced cognitive reflection tendencies seen in those who display lower DE (14). Notably, a distinct exploratory strategy – ‘random’ exploration (i.e., where choices become less reward-driven as a means of gaining information) – has not shown such associations.

Past research therefore suggests that negative affect may selectively reduce DE, consistent with maintained avoidance. Yet, these studies are largely correlational and have tended to focus on trait anxiety, and they have primarily investigated sub-clinical symptoms in community samples. There are also some reasons to expect relationships in the opposite direction. Namely, the cognitive aspects of anxiety involve elevated uncertainty and worry, which could promote over-exploration. Indeed, intolerance of uncertainty could drive over-exploration as a means of continually attempting to reduce it. This is consistent with some work instead suggesting greater DE in those with higher trait anxiety (29, 30) and increased exploratory behavior in depression (30, 31). Combined with the work reviewed above, this suggests the possibility that state or somatic anxiety might reduce exploration, while uncertainty- or worry-related trait or cognitive anxiety could instead promote it.

Here we sought to build on this previous work by manipulating state anxiety with a somatic anxiety induction while individuals performed an explore-exploit task. By comparing exploration and learning with vs. without this induction, while also gathering information about trait anxiety and other clinical, cognitive, and affective dimensions, we aimed to disentangle the role of state- and trait-related anxiety. By doing this in both healthy comparison (i.e., low-negative affect) and clinical (i.e., high-negative affect) groups, we also sought to clarify potential differences between subclinical variation in anxiety and depression and that associated with psychopathology. We hypothesized that state anxiety induction would reduce DE. This would follow from the possibility that anxious somatic (high arousal) states plausibly disrupt reflective cognition and attention to uncertainty (e.g., see 15, 24, 25, 26). Given the mixed results described above with respect to trait anxiety, we did not have a confident directional hypothesis regarding group differences. As secondary aims, we also sought to reproduce the aforementioned relationships found between DE and both cognitive reflectiveness (14) and early adversity (16, 17), as these associations could offer additional explanatory power if present in our sample.

## Methods

### Participants

Data were collected as part of a larger, multi-visit study to investigate the cognitive and neural correlates of psychiatric disorders at the Laureate Institute for Brain Research (LIBR), with participants recruited from the community in and surrounding Tulsa, OK, USA. Clinical diagnoses were assessed by a licensed clinician according to the Mini International Neuropsychiatric Interview 7 (MINI; (32)). Healthy comparisons (HCs) were not previously diagnosed with, or treated for, any mental health disorder and had a score of < 8 on the Overall Anxiety Severity and Impairment Scale (OASIS; (33)). The clinical group consisted of individuals with one or more affective disorders displaying elevated anxiety symptoms (iADs). Due to high comorbidity rates between depression and anxiety diagnoses (34, 35), those in this group were permitted to have either a current or lifetime diagnosis of anxiety and/or depressive disorders and were required to have a score > 8 on the OASIS in a pre-visit screening. For a full comorbidity breakdown in iADs, see **Supplementary Table S1**. Any iADs who were taking psychiatric medications (e.g., SSRIs, SNRIs, etc.) were required to have a stable dosage for at least 6 weeks prior to study entry. Participants were asked to abstain from other medications (e.g., benzodiazepines), marijuana, and alcohol in the 48 hours preceding participation and were required to pass a drug panel during their study visit. Antipsychotic and stimulant medications were not allowed (a detailed list of included/excluded medications is provided in **Supplementary Materials**). The following diagnoses were also not permitted to minimize confounding influences on primary outcome measures: bipolar disorder, personality disorders, substance use disorders, eating disorders, schizophrenia, or obsessive-compulsive disorder.

Recruitment aimed to match participants in the two groups by age, sex, and education level.

Seven individuals withdrew from participation due to discomfort with the anxiety induction paradigm (2 HCs, 5 iADs, all female; details in **Supplementary Materials**), leading to a final sample of 119 participants: 58 HCs, 61 iADs (see **Table 1**). A post-hoc power analysis (using the *wp.rmanova* function in the *WebPower* package in R (36)) indicated that this sample size would afford 80% power to detect a medium effect size of 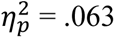 for the effect of anxiety induction on task behavior. While no prior studies are available (to our knowledge) reporting effects of breathing-based anxiety induction on decision-making, we note that some previous work has assessed effects of stress induction on potentially related cognitive tests (e.g., executive function batteries; 37). Here, effect sizes for stress-induced changes in test scores have tended to be large (Cohen’s *d*>.88). Thus, results of the above power analysis for the present study suggest a sufficiently low false negative rate, assuming similarity to effects reported in this prior work.

**Table 1.** Demographic and clinical characteristics (mean and SD). *This measure was collected as part of the larger study at an earlier date (35 days prior to participation on average).

| Measure | Healthy Comparisons<br>(N=58) | Affective Disorders<br>(N=61) | Statistic |
| --- | --- | --- | --- |
| Age | 35.41 (13.08) | 33.44 (10.97) | $t(117) = 0.89, p = .374$ |
| Sex | 72% Female | 75% Female | $\chi^2(1) = 0.03, p = .870$ |
| PHQ | 1.66 (2.27) | 9.46 (5.15) | $t(117) = 10.60, p < .001$ |
| OASIS | 1.07 (1.52) | 8.41 (3.02) | $t(117) = 16.62, p < .001$ |
| QIDS | 2.17 (1.77) | 8.77 (4.01) | $t(117) = 11.52, p < .001$ |
| STAI Trait | 29.17 (7.4) | 51.87 (9.06) | $t(117) = 14.92, p < .001$ |
| CTQ Total* | 34.44 (10.36) | 48.07 (16.19) | $t(112) = 5.31, p < .001$ |
| Physical Neglect | 6.18 (2.14) | 8.41 (3.79) | $t(112) = 3.82, p < .001$ |
| Physical Abuse | 6.67 (2.33) | 7.95 (3.26) | $t(112) = 2.39, p = .019$ |
| Emotional Neglect | 8.76 (3.88) | 13.03 (5.02) | $t(112) = 5.05, p < .001$ |
| Emotional Abuse | 6.89 (2.98) | 10.59 (4.59) | $t(112) = 5.07, p < .001$ |
| Sexual Abuse | 5.93 (3.05) | 8.08 (5.01) | $t(112) = 2.75, p = .007$ |
| CRT | 3.33 (2.18) | 2.18 (1.93) | $t(117) = 3.05, p = .003$ |

**Note.** While OASIS scores at screening were required to be $\geq 8$ for inclusion in the Affective Disorders group, values shown here are at the later date of study participation. As OASIS scores are rated with respect to the prior week, some scores represented here were $< 8$ in this group. Legend: PHQ = Patient Health Questionnaire-9 (depressive symptoms); OASIS = Overall Anxiety Severity and Impairment Scale; STAI = State-Trait Anxiety Inventory, CTQ = Childhood Trauma Questionnaire; CRT = Cognitive Reflection Test (scores correspond to number of correct answers).

It is important to note here that data from the healthy sample in this study has also been used for comparison to a different clinical population (methamphetamine use disorders) (38). Here, we focus on differences between this group and iADs.

### Measures

Primary measures were chosen to assess state and trait anxiety (State-Trait Anxiety Inventory [STAI]; (39)) and to account for clinical symptoms of depression (Patient-Health Questionnaire [PHQ]; (40)). Our secondary aim of replicating prior work motivated inclusion of measures to test links between DE and both childhood adversity (Childhood Trauma Questionnaire [CTQ]; (41)) and cognitive reflectiveness (Cognitive Reflection Test [CRT]; (14, 16, 17)) – the tendency to ‘think things through’ before responding based on intuition. Detailed descriptions of each measure are included in **Supplementary Materials**. Descriptive information for all participants is shown in **Table 1**, as well as preliminary group comparisons. Additional sample demographics are included in **Supplementary Table S2**.

### Somatic Anxiety Induction and Sensitivity Assessment

To manipulate state anxiety, we utilized a previously established interoceptive (breathing-based) anxiety induction paradigm (42–44). In this paradigm, participants are asked to breathe through a silicon mask attached to a valve (**Figure 1a**) that allows application of different levels of inspiratory resistance, creating air hunger-related sensations that induce anxiety. To provide participants a chance to familiarize themselves with the paradigm, and assess baseline sensitivity, participants were first exposed to a series of resistances in ascending order (0, 10, 20, 40, 60, and 80 cmH2O/L/sec) applied for 60 seconds each. Participants were asked to rate their anxiety level after each exposure from 0 (“no anxiety”) to 10 (“maximum possible anxiety”). Other questions relating to subjective breathing difficulty, arousal, and other affective states were also presented (see **Supplementary Materials**).

**Figure 1.**
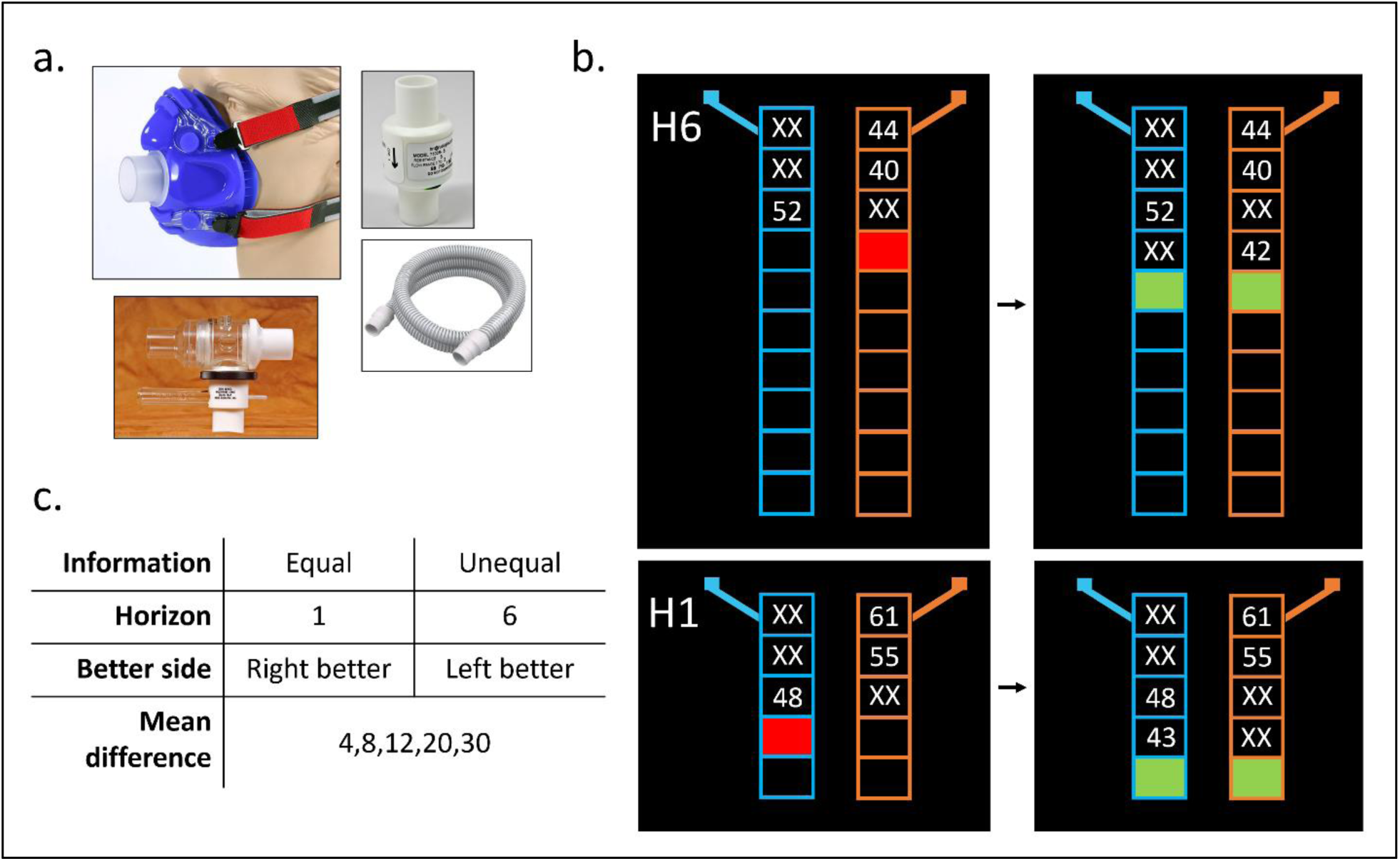
Anxiety induction apparatus and behavioral task. a) Equipment used for anxiety induction including silicon mask with adjustable straps and single breathing port, and an example resistor used to create resistance on inhalation (equipment manufactured by Hans Rudolph inc.). b) Graphical depiction of the Horizon Task. Shown are the two horizon conditions (H1 and H6) at the last forced choice (red box) and the first free choice (green boxes). The H1 example also shows an equal information trial (2 outcomes for each choice), while the H6 example shows an unequal information trial (3 forced choices on the right, 1 on the left) designed to promote directed exploration (i.e., choosing the side for which only one outcome has been observed). c) Table outlining each of the trial type combinations, counterbalanced across the task.

This induction was later applied using a moderate resistance level (40 cmH2O/L/sec) during performance of one run of the behavioral task. This level of resistance was chosen because of prior work demonstrating its effectiveness and feasibility (45); we also confirmed that it induced moderate but tolerable (roughly 5 out of 10) levels of anxiety within our clinical sample in the initial sensitivity assessment (**Supplementary Figure S2**).

### Behavioral Task

Participants completed two runs of the Horizon Task (46), a previously validated task used to measure explore-exploit decision-making. Each task run consists of 80 independent games. In each game, players are presented with two new options (slot machines) and asked to repeatedly choose between them. They are told that the average reward values for options on one game are unrelated to those on other games. Half of the games include 5 sequential choices, while the other half include 10 sequential choices. The first four choices in all games are “forced”, meaning the player is told which option to choose (see **Figure 1b**). The remaining choices are “free”, where either option can be chosen. Trials with one free choice (H1) and those with six free choices (H6) appear an equal number of times throughout the task.

Forced-choice patterns create two different information conditions: *equal*, in which each option is chosen twice; and *unequal*, in which one option is chosen three times and the other is chosen only one time. The goal of the task is to maximize the number of points won by repeatedly choosing the option expected to provide higher point values on average. For each option, point values were sampled (rounded to the nearest integer) from a Gaussian distribution with a standard deviation of 8. One of the options in each game always had a mean reward value of either 40 or 60. Then, for each game, the other option had a mean point value difference from this first option of +/- 4, 8, 12, 20, or 30. The same pseudorandom sequences of forced-choice outcomes were shown to each participant, consistent with the generative means. All game dimensions (length, information type, mean difference, and better/worse side) were counterbalanced across the task (**Figure 1c**). For maximal comparability, each of the two task runs was kept identical in terms of mean differences, forced-choice outcomes, and the associated game order.

Participants wore the mask for each task run. In the *no-resistance run*, resistance was not added (0 cmH2O/L/sec). In the *resistance run*, a moderate resistance (40 cmH2O/L/sec) was consistently applied throughout the run. The order was counterbalanced across participants in each group. The first six participants played a version of the task with different outcome values than the rest in a small number of games (but still sampled from the same underlying distributions). To ensure this did not confound results below, this difference in task version was evaluated as a potential covariate.

### Computational Model and Model Fitting

Here, we followed the same modeling and parameter estimation approach used in previous studies with this task (e.g., 47, 48). This approach is presented in detail in **Supplementary Materials**. Briefly, the model assumes participants start each game by assigning an initial expected reward value to the two new options presented (here fixed at a neutral value of 50). These expected reward values are then updated after each forced-choice outcome based on the resulting prediction error, moderated by an evolving (uncertainty-dependent) learning rate.

Finally, the resulting expected reward value differences between options are used to make the first free choice, moderated by two parameters (i.e., information bonus and decision noise) promoting different forms of exploration. Specific definitions of each computational parameter are provided in **Table 2**. As each game in the task is independent (i.e., with two new options presented, along with a new set of forced-choice outcomes), the model assumes no learning carries over between games or task runs. Independent parameter values were therefore estimated for each task run for each participant. This estimation step was carried out simultaneously for both groups and conditions with an established hierarchical fitting procedure using a Markov Chain Monte Carlo method (see **Supplementary Materials**).

**Table 2.** Descriptions of model parameters.

| Model Parameter | General Description |
| --- | --- |
| Information Bonus (IB) | IB parameters (separated by horizon condition to yield IB1 and IB6) quantify an individual's tendency to choose the option that maximizes information gain. This parameter only affects decisions in the unequal information condition, where selecting the option that was chosen only once during the forced choices provides more information about its average reward value. Directed exploration (DE) is derived from these parameters (IB6 minus IB1). This is because information gain in H1 cannot guide future choices, while information gain in H6 can guide future choices. Thus, DE can be seen as the tendency to increase information-seeking when it would be useful. |
| Decision Noise (DN) | The DN parameters (separated by horizon condition to yield DN1 and DN6) quantify an individual's tendency to choose the option with a lower observed mean reward. Random exploration (RE) is derived by subtracting DN1 from DN6 in the equal information condition. Thus, RE can be seen as the tendency to make less reward-sensitive choices when information gain would be useful. |
| Spatial Bias | These parameters (one per horizon/information condition) account for the possibility that an individual prefers to choose one option over the other simply because of which side it is on. |
| Initial Learning Rate ($\alpha_0$) | The initial value of the learning rate before making the first forced choice. |
| Asymptotic Learning Rate ($\alpha_\infty$) | The learning rate an individual would converge to if the game were played indefinitely (i.e., when uncertainty about mean reward values would be minimal). |

### Statistical Analyses

#### Resistance sensitivity

To verify that the anxiety induction was effective, we examined affective responses to each resistance level in the initial exposure protocol. To this end, we estimated linear mixed effects models (LMEs) with self-reported anxiety (i.e., the single anxiety question on a scale from 0 to 10) as the outcome variable and resistance level and group as regressors (and their interaction).

#### Primary model-based analyses

Before addressing our primary hypotheses, we checked for outliers using an iterative Grubbs method (threshold: *p*<.01; using *grubbs.test* from the *outliers* package in R (49)). This resulted in one data point being removed for IB1 (in the resistance condition), two for DN6 in the equal information condition (one in each resistance condition), and two for RE (one in each resistance condition). Then, we estimated LMEs that included group, resistance condition, and their interaction as sum-coded regressors for each model parameter (HCs=-1, iADs=1; no-resistance=- 1, resistance=1). Given that the two learning rates were significantly positively correlated in both task runs (*rs*>.34, *p*<.001), models with 𝜶_∞_ as the outcome variable were also tested when accounting for 𝜶_𝟎_ values to assess whether unique relationships with asymptotic learning rate were present even after accounting for its shared variance with initial learning rate. To rule out possible alternative explanations, we also re-ran these models including covariates to confirm that any group or resistance effects could not be explained by age, sex (Male=-1, Female=1), or task version (new=-1, old=1).

We then assessed continuous relationships between computational parameters and affective symptoms. In each group separately, we tested linear models of DE in each resistance condition using PHQ scores and STAI Trait scores as regressors (also accounting for age). However, given the high inter-correlations between PHQ and STAI, problematic multicollinearity issues were possible. To evaluate this, we calculated variance inflation factors (VIFs; using the *ols_vif_tol* function from the *olsrr* package in R (50)) for each model and, in cases where VIFs surpassed a threshold of 4, we planned to use ridge regressions designed to minimize problematic multicollinearity effects (using the *linearRidge* function from the *ridge* package (51)).

#### Model-free task performance

To better interpret observed effects on task behavior and assess relationships between task performance and model parameters, we performed additional supportive analyses examining overall accuracy on free choices (as measured by the number of times participants chose the option with the higher underlying mean reward). We first restricted analyses to first free choice and estimated an LME with accuracy as the outcome variable and horizon, information condition, resistance, and group as regressors. We also included possible three-way interactions between group, horizon, and information condition, and between group, horizon, and resistance (and associated two-way interactions). This allowed us to explore whether groups might show greater differences depending on horizon, information condition, or anxiety induction. As commonly observed in this task, we expected first free choice accuracy in H1 trials would be higher than in H6 trials (i.e., reflecting random exploration).

A subsequent model was run associating accuracy with free choice trials in H6 (choices 5-10). We also tested three-way interactions between group, free choice number, and information condition, and between group, free choice number, and resistance (and associated two-way interactions) to explore whether groups might differ in the slope of accuracy improvement over time depending on information condition or anxiety induction.

To evaluate whether some parameter values might be considered more optimal than others, we then tested LMEs associating parameters with accuracy on H6 free choice trials, excluding the first free choice to which model parameters were fit. As IB is only calculated in the unequal information condition, we only included accuracy on trials for this condition in these models, with resistance condition, group, free choice number, IB for H6 (IB6), and the interaction between free choice number and IB6 as regressors. Analogous models were used to test relationships between accuracy and the other model parameters within the relevant trial types (i.e., decision noise for H6 with equal information condition accuracy; learning rates with accuracy for both information conditions).

#### Secondary replication analyses

Additional LMEs were run to accomplish our secondary aim of replicating prior relationships found between DE and both early adversity (16, 17) and cognitive reflectiveness (14). First, separate LMEs were run on DE based on each subscale of the CTQ, while also including group, resistance condition, and their interaction as regressors (while controlling for known effects of age). Analogous LMEs were run instead using CRT scores as regressors.

## Results

### Model Validation

We first assessed parameter recoverability. Specifically, participants’ fitted parameter values were used to simulate behavior under the model, and this simulated behavior was then used for parameter estimation. The correlation between generative and estimated parameter values in these simulations provides a measure of parameter recoverability. All correlations were significant and positive (*rs*>.52, *ps<.*001), suggesting moderate-to-good recoverability (see **Supplementary Figure S1**); however, we note that IB1 was somewhat lower in recoverability than the other parameters. This was likely because observed values for this parameter tend to show a restricted range near zero, as the H1 condition is designed to minimize the value of information. Recoverability for DE itself was notably higher (*r*=.78) and correlated more strongly with IB6 values (*r*=.78), suggesting the IB1 estimates acted more like stable floor values.

We then confirmed that parameter estimates correlated with descriptive measures of task behavior in expected directions (e.g., percentage of trials in which the high-information and low-mean option was chosen, points won). These results were all in expected directions (detailed in **Supplementary Materials**). In particular, IB parameters were positively associated with the percentage of trials in which the high-information option was chosen (*rs*>.86, *ps*<.001 for both resistance conditions), and DN parameters were positively associated with the percentage of trials in which the low-mean option was chosen (*rs*>.29, *ps*<.002). Slower learning rates were also associated with a greater number of trials in which the low-mean option was chosen (*rs*<- .43, *ps*<.001) and worse task performance in general (i.e., fewer points won on the task; *rs*>.45, *ps*<.001).

### The Breathing Resistance Protocol Successfully Induced Anxiety During Task Performance

Anxiety ratings during the initial breathing resistance sensitivity protocol are shown in **Supplementary Figure S2**. Briefly, anxiety increased as resistance level increased, anxiety ratings were higher in iADs than HCs, and iADs showed greater increases in anxiety compared to HCs as resistance level increased. LMEs for self-reported anxiety scores at baseline and during both task runs showed the same pattern (**Figure 2**). See **Supplementary Materials** for full results.

**Figure 2.**
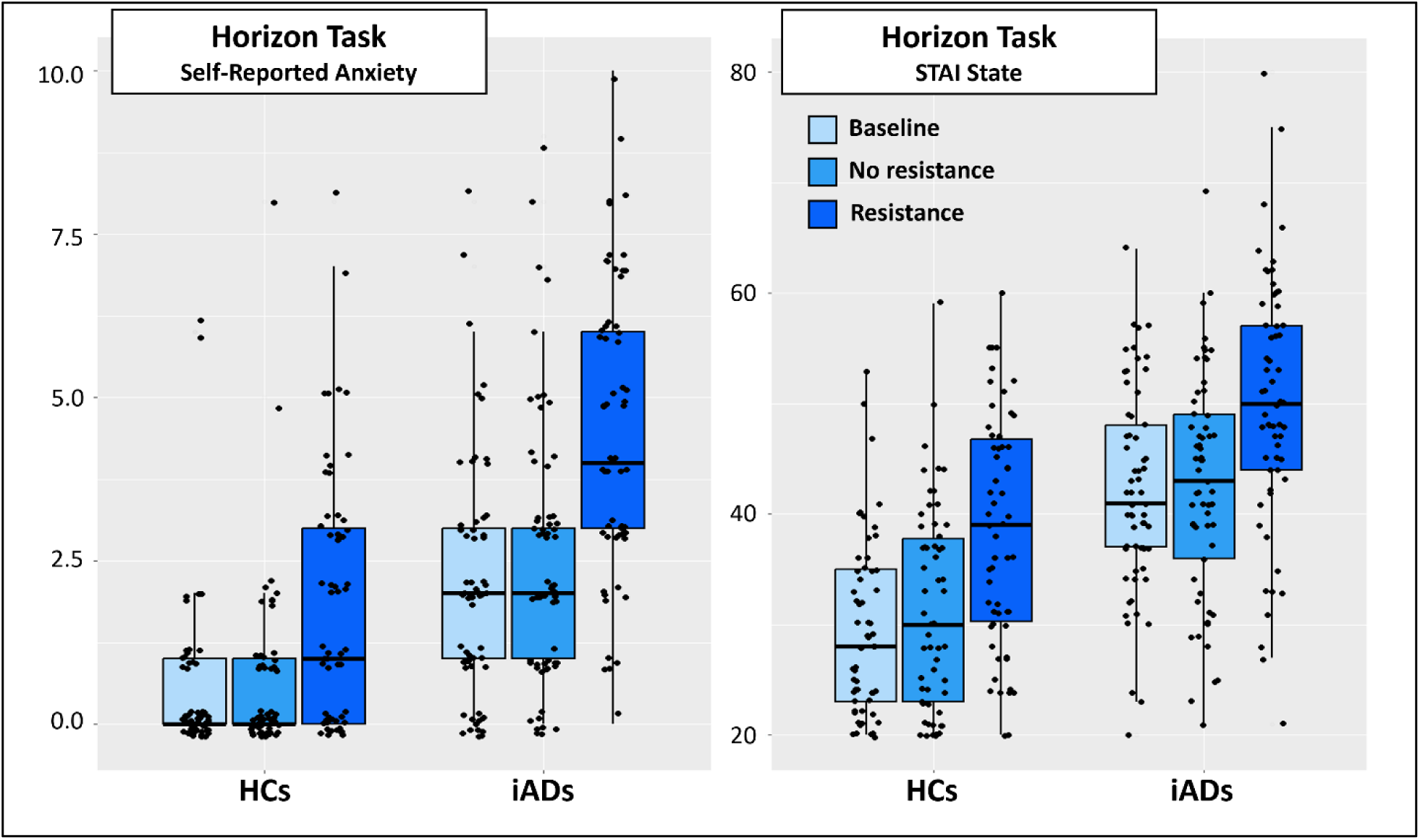
Self-reported anxiety and STAI State at baseline and during each run of the Horizon Task (i.e., with and without the added breathing resistance of 40 cmH2O/L/sec). Results showed greater anxiety in iADs, greater anxiety with higher resistance level, and greater increases in anxiety in iADs than HCs when resistance was added. Boxplots show median and quartile values along with individual datapoints.

### Directed Exploration was Reduced by State Anxiety Induction in Healthy Participants but not in those with Affective Disorders

Plots depicting parameter values by group and condition are shown in **Figure 3**. Descriptive statistics for model parameters are reported in **Supplementary Table S4.** In an initial LME for DE, there was a significant *Group x Resistance Condition* interaction (*F*(1,117)=3.97, *p*=.049, 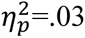), such that HCs and iADs showed similar DE in the no-resistance condition, DE in HCs was lower in the resistance condition (estimated marginal mean [EMM]= 4.28) than in the no-resistance condition (EMM=5.27; *t*(117)=-2.52, *p*=.013), and DE in iADs did not change between conditions (*t*(117)=0.27, *p*=.786). Further exploration suggested this was driven primarily by differences in IB6 values within the resistance condition (see top-right panel of **Figure 3**). Main effects of group and resistance were nonsignificant. In a subsequent model accounting for age, sex, and task version, this interaction remained significant (*F*(1,117)=3.97, *p*=.049, 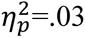). There was also a negative association with age (b=-0.06; *F*(1,114)=5.65, *p*=.019, 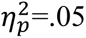).

**Figure 3.**
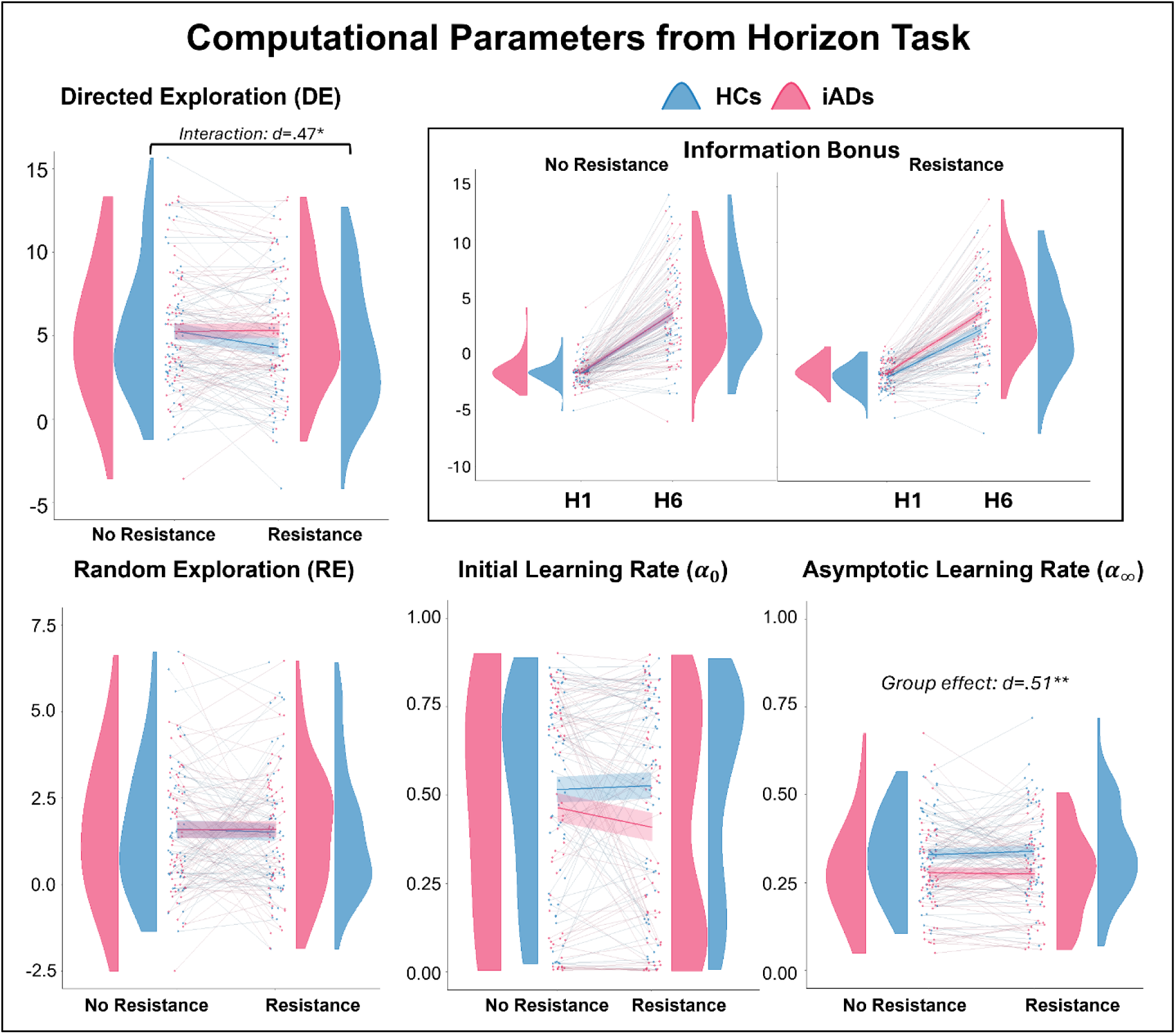
Individual data points and density plots by group and resistance condition for each parameter. In HCs, the anxiety induction led to reduced levels of DE. In contrast, iADs were insensitive to this effect and showed stable DE values in both conditions. For illustrative purposes, the individual information bonus (IB) values underlying DE for each horizon condition (H1 and H6) are shown in the top-right (recall that DE corresponds to the difference in information bonuses between horizon conditions; DE = IB6 – IB1). As can be seen, only IB6 values in the resistance condition show differences between groups. RE did not differ by group or condition (bottom left). Learning rates tended to show lower values in iADs than HCs across conditions. As visible in the lower middle panel, 𝜶_𝟎_ showed a bimodal distribution, which motivated the cluster-based analysis approach described in the text. Asymptotic learning rates (𝜶_∞_) were significantly lower in iADs than HCs in both conditions (bottom right). Note that stars and Cohen’s *d* effect sizes correspond to the interaction between group and resistance for DE (upper left) and for the group difference in 𝜶_∞_across resistance conditions (bottom right) in post-hoc comparisons.

No significant effects were found in LMEs for RE (*Fs*<3.83, *ps*>.053).

Because of bimodality in the distributions for 𝜶_𝟎_ (see **Figure 3**), we used *k*-means clustering (52) across resistance conditions to divide participants into groups with high vs. low values and treated this as a 2-level categorical variable (low=-1; high=1). In logistic regression models using membership in the high vs. low 𝜶_𝟎_**-**value group as the outcome variable (using *glmer* in the *lme4* package; (53)), there were no significant effects before or after adding covariates (*zs*<|1.53|, *ps*>.125). Complementary nonparametric models treating 𝜶_𝟎_as a continuous outcome variable (using the *Rfit* package in R (54)) revealed a nonsignificant trend (b=-0.03, *p*=.052) suggesting potentially higher 𝜶_𝟎_ values in HCs than iADs. A main effect of sex (b=-0.06, *p*=.003) was also found, indicating higher 𝜶_𝟎_ values in male participants.

The LME for 𝜶_∞_ (accounting for 𝜶_𝟎_) revealed a significant main effect of group (*F*(1,117)=7.72, *p*=.006, 𝜂_p_^2^=.07), such that HCs (EMM=.33) had higher learning rates than iADs (EMM=.28, *t*(117)=-2.78, *p*=.006). The expected positive association with 𝜶_𝟎_ was also observed (*F*(1,199)=16.31, *p*<.001, 𝜂_p_^2^=.08; b=.11). Both effects remained significant when accounting for covariates, and no additional effects were observed (*Fs*<1.74, *ps*>.189).

We also evaluated within-group associations with symptom severity to explore potentially differential effects of depression and anxiety. We noted that PHQ and STAI Trait scores were highly correlated (*rs*=.86; see **Supplementary Figure S4**); yet, all VIFs<4, suggesting multicollinearity did not threaten the validity of these models. Within HCs, a linear regression with DE in the no-resistance condition as the outcome variable revealed no significant relationships (*Fs*<1.95, *ps*>.169). However, there was a significant positive effect of STAI Trait in the resistance condition (b=.193, *F*(1,55)=4.17, *p*=.046) that became marginal after adding covariates (b=.167, *F*(1,53)=2.93, *p*=.093). PHQ showed no associations with and without covariates. In iADs, no significant effects were observed in equivalent analyses. These analyses were also carried out for other model parameters (detailed in **Supplementary Results**). Briefly, these suggested 1) opposing relationships with 𝜶_∞_for depression (negative) and anxiety (positive) symptoms in HCs in the no-resistance condition that flipped in the resistance condition (i.e., positive relationship with depression and negative with anxiety), 2) potential negative associations between trait anxiety and 𝜶_𝟎_in HCs in the resistance condition, and 3) a possible relationship between RE and depression (negative) for iADs in the no-resistance condition.

### Higher Levels of Directed and Random Exploration were Associated with Greater Task Performance

In LMEs for H6 accuracy (on choices 6-10) using each model parameter, we found interactions between free choice number and IB6, DN6, and 𝜶_𝟎_(**Table 3**). Specifically, the interaction between IB6 and free choice number indicated that, in unequal information trials, those with higher IB6 values showed steeper increases in accuracy from early to later choices. Similar results were found for DN6 when examining accuracy on equal information trials and for 𝜶_𝟎_on unequal information trials.

**Table 3.** Models of free choice accuracy (separated by information condition) testing associations with model parameters.

|  | Parameter | Resistance | Group | Free Choice Number | Parameter*Free Choice |
| --- | --- | --- | --- | --- | --- |
| **IB6**<br>(unequal IC) | $F(1,987)=1.71$,<br>$p=.191$ | $F(1,1078)=.14$,<br>$p=.711$ | $F(1,117)=1.63$,<br>$p=.204$ | $F(1,1067)=91.96$,<br>$p<.001$; $b=0.017$ | $F(1,1067)=7.35$,<br>$p=.007$; $b=0.001$ |
| **DN6**<br>(equal IC) | $F(1,1173)=3.16$,<br>$p=.076$ | $F(1,1059)=2.40$,<br>$p=.122$ | $F(1,117)=5.69$,<br>$p=.019$ | $F(1,1057)=113.00$,<br>$p<.001$; $b=0.013$ | $F(1,1057)=6.28$,<br>$p=.012$; $b=0.003$ |
| **$\alpha_0$ Cluster**<br>(equal IC) | $F(1,1170)=2.03$,<br>$p=.154$; $b=-.014$ | $F(1,1067)=256$,<br>$p=.110$ | $F(1,117)=5.67$,<br>$p=.019$ | $F(1,1067)=114.48$,<br>$p<.001$; $b=.025$ | $F(1,1067)=1.66$,<br>$p=.198$ |
| **$\alpha_0$ Cluster**<br>(unequal IC) | $F(1,1159)=5.63$,<br>$p=.018$ | $F(1,1067)=.01$,<br>$p=.919$ | $F(1,117)=1.67$,<br>$p=.199$ | $F(1,1067)=91.67$,<br>$p<.001$;<br>$b=0.016$ | $F(1,1067)=11.34$,<br>$p=.001$; $b=0.016$ |
| **$\alpha_\infty$**<br>(equal IC) | $F(1,1172)=.65$,<br>$p=.419$ | $F(1,1067)=2.37$,<br>$p=.124$ | $F(1,119)=5.43$,<br>$p=.022$ | $F(1,1067)=114.59$,<br>$p<.001$; $b=.03$ | $F(1,1067)=.66$,<br>$p=.418$ |
| **$\alpha_\infty$**<br>(unequal IC) | $F(1,1165)=.42$,<br>$p=.518$ | $F(1,1067)=.036$,<br>$p=.850$ | $F(1,118)=1.69$,<br>$p=.196$ | $F(1,1067)=91.13$,<br>$p<.001$; $b=0.033$ | $F(1,1067)=2.97$,<br>$p=.085$ |

**Note.** For each significant effect of group, contrasts indicated that accuracy in iADs (EMM=0.80) was lower than in HCs (EMM=0.85; $p<.05$ ). The models of accuracy including $\alpha_\infty$ also showed positive effects of $\alpha_0$ ( $F_s \geq 8.83$ , $p_s \leq .003$ , $b_s \geq 0.055$ ). IC=information condition.

We also tested an analogous model of free choice accuracy using DE as a regressor and saw comparable results to those seen for IB6 (i.e., main effect of free choice number and a significant interaction with DE; *Fs*>6.11, *ps*<.014). Similarly, results when using RE in a model of equal information accuracy corroborated those found for decision noise (i.e., main effect of free choice: *F*(1,1062)=114.45, *p*<.001; *RE x free choice*: *F*(1,1062)=8.92, *p*=.003; main effect of group: *F*(1,117)=5.60, *p*=.020).

### Task Performance was Reduced in Affective Disorders in Specific Conditions

Secondary analyses of free choice accuracy indicated high performance across both groups (HCs: M=.84, SD=.13; iADs: M=.80, SD=.16; see **Supplementary Figure S5**). LMEs for first free choice accuracy (**Supplementary Table S5**) revealed significant effects of horizon (greater accuracy in H1; *F*(1,823)=278.81, *p*<.001) and information condition (greater accuracy in equal information trials; *F*(1,823)=72.08, *p*<.001). The *Group x Horizon x Information Condition* interaction was also significant (*F*(1,823)=4.18, *p*=.041), indicating that HCs had significantly greater accuracy than iADs in H1/equal information trials and in H6/unequal information trials only (see post-hoc contrasts). Subsequent LMEs for accuracy across free choices in H6 showed significant effects of choice number (greater accuracy in later choices, as expected; *F*(1,2727)=201.30, *p*<.001), information condition (greater accuracy in equal information trials; *F*(1,2727)=51.37, *p*<.001), and a *Group x Information Condition* interaction (greater accuracy in HCs only in the equal information condition; *F*(1,2727)=8.93, *p*=.003).

### Replication Analyses Confirmed Associations with Early Adversity and Cognitive Reflectiveness

CTQ scores were greater in iADs than HCs (*t*(112)=5.31, *p*<.001). In separate LMEs for DE with each subscale of the CTQ, group, resistance condition, and their interaction as regressors (controlling for age), and after correcting for multiple comparisons (*p*=.01), there was a significant negative association with physical abuse (b=-.76; *F*(1,109)=6.96, *p*=.010, 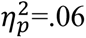) such that exploration was lower in those who experienced greater physical abuse. This remained significant in iADs alone (b=-1.05; *F*(1,56)=7.27, *p*=.009, 𝜂_p_^2^=.11). No effects were observed for any other parameter or CTQ subscale.

CRT scores were lower in iADs than HCs (*t*(117)=3.05*, p*=.003). In an LME assessing the effect of CRT on DE across all participants, there was a significant positive association (b=.80; *F*(1,114)=6.46, *p*=.012, 𝜂_p_^2^=.05). The effect of CRT was directionally the same in iADs alone (and stronger in effect size), but only marginally significant due to smaller sample size (b=.77; *F*(1,58)=3.89, *p*=.053, 𝜂_p_^2^=.06). There were no significant effects of CRT on RE or 𝜶_∞_ in either the full sample or in the clinical group alone (*ps*>.053). Higher CRT scores were associated with membership to the cluster with high 𝜶_𝟎_ values in the full sample (z=-2.72, *p*=.007; log-odds=- .85, CI=[-1.46,-0.24]), but not in iADs alone (z=-1.61, *p*=.108; log-odds=-.77, CI=[-1.71,0.17])).

Complementary nonparametric models using 𝜶_𝟎_ as a continuous outcome variable also revealed a significant positive association with CRT scores in both the full sample (b=0.08, *p*<.001) and in iADs alone (b=0.07, *p*=.003).

## Discussion

In this study, we compared decision-making behavior on an explore-exploit task in participants with and without clinically significant levels of anxiety and depression. A somatic (interoceptive) anxiety induction was used to dissociate the influences of state vs. trait anxiety. We hypothesized that state anxiety induction would reduce directed exploration (DE) as a potential mechanism promoting avoidance behavior. Results were mixed, offering only partial support for this hypothesis. First, as predicted, DE was reduced by the anxiety induction in HCs, suggesting reduced reflection on uncertainty. This is consistent with other correlational work in non-clinical samples linking anxiety to reduced DE (14, 15), as well as work showing that stress reduces exploration (21–23). Notably, while choice accuracy did not differ between conditions, higher levels of DE were associated with steeper improvements in task performance over time, suggesting reductions in DE during the anxiety induction could be viewed as maladaptive.

In contrast, DE in iADs remained stable between conditions. One interpretation of this result is that, as iADs experience greater state anxiety on a regular basis, they may have developed compensatory strategies that led them to be insensitive to this effect. This is consistent with other work suggesting compensatory processes in anxious individuals that allow them to maintain good performance (e.g., see 55, 56). On the other hand, these stable DE levels across conditions also suggest iADs had greater uncertainty during the anxiety induction about the average reward value of each choice in the task, or greater sensitivity to the difference in their uncertainty between options. This appears consistent with a few recent studies suggesting greater DE in those with higher levels of cognitive anxiety or worry (e.g., 29, 30). .

We also found that iADs showed slower learning rates across conditions, which may offer some additional explanatory insights. First, it is worth noting that these learning rates were also positively associated with task performance. This therefore suggests that slower learning may have contributed to worse performance in iADs. Here, slower learning implies that beliefs remained closer to uninformative prior values; thus, confidence in the better choice would increase more slowly (similar to less flexible learning rates previously associated with higher anxiety; (5)). Within this task, slower learning rates might thus support a type of persistent uncertainty about the estimated values of each bandit, with suboptimal effects on decision-making. Interpreting this result in light of previous findings is subtle, however, as faster learning rates are theoretically linked to greater uncertainty, and prior work has linked anxiety to both faster learning (6) and a greater tendency to infer changes in context (8). On the other hand, the type of uncertainty in these studies pertains to volatility, or how frequently environmental contingencies are expected to change. Thus, if anxious individuals believe the world is ever-changing, then learning rates should be high.

However, if “uncertainty” instead pertains to the estimated stochasticity of (i.e., noise in) the mapping from underlying states to observations, learning rate should instead be low, so as not to overfit beliefs to random outcomes (57). Thus, in the present task, slower learning in iADs could represent greater uncertainty about the informativeness of each outcome when inferring the underlying reward mean. This could offer a complementary means by which uncertainty is maintained in negative affect, consistent with the previous findings reviewed above. It could also relate to another recent study showing that individuals who experienced greater early adversity, itself a predictor of subsequent affective disorders (58–60), also showed slower learning rates (16). It should be kept in mind, however, that our sample includes individuals with affective disorders and used an anxiety induction, while most prior work has examined sub-clinical levels of anxiety and depression using correlational approaches. Thus, some results between studies may not be fully comparable. This was suggested by our follow-up results in which DE in HCs showed positive associations with trait anxiety in the resistance condition (i.e., even accounting for age and depression), while no such relationships were found in the clinical sample. These results support specific associations with trait anxiety, but also highlight how prior results in non-clinical samples (reviewed in 61) may not generalize to clinical samples (or perhaps suggest ceiling effects).

In line with our secondary aims, we were also able to successfully replicate prior results showing higher DE in those with greater cognitive reflectiveness (14) and lower DE in those who had experienced greater childhood adversity (i.e., physical abuse; 16, 17). These findings suggest that less reflection on uncertainty and greater exposure to unpredictable/harsh early environments could each contribute to clinical differences seen here in exploration and learning. It is also worth noting that cognitive reflectiveness has been shown to improve with training (62–64).

Future studies could therefore examine whether improving reflectiveness also optimizes uncertainty estimation and/or reduces affective symptoms. Another consideration is that multiple previous studies have shown strong relationships between physical abuse in childhood and later avoidance behavior (65, 66). Our results thus suggest differences in DE could contribute to this avoidance behavior in those recovering from childhood trauma, highlighting DE as a possible treatment target.

There are important limitations to consider. First, state anxiety levels were higher in iADs than HCs in both the no-resistance and resistance conditions. Thus, effects of state anxiety level may not be fully comparable between groups (particularly if non-linear relationships are present).

Additionally, the sample size was only moderate and may not have afforded sufficient power to detect some effects within the clinical group alone. Future research will also be needed to see whether results generalize to other explore-exploit tasks as well as tasks designed to distinguish learning rates in relation to volatility vs. stochasticity (57).

## Conclusions

The results of this study suggest that somatic anxiety induction reduces directed exploration in healthy individuals, while those with affective disorders may be insensitive to this effect – possibly due to learned compensatory strategies (55, 56). They also suggest those with affective disorders display slower learning rates generally. Results further confirm lower levels of directed exploration in those displaying less cognitive reflection and in those who have experienced greater early adversity. These findings highlight potential computational mechanisms underlying the influence of anxiety on uncertainty estimation and maladaptive avoidance. If confirmed in future work, this could suggest potential benefits of treatments aimed at optimizing reflection on uncertainty, levels of information-seeking, and belief testing in relation to current affective states, as well as adjusting beliefs about the reliability of new experiences in revising expectations.

## Author contributions

NL and CAL took the lead in data analysis and writing the manuscript. RS designed the study, planned and oversaw analyses, and guided manuscript writing. NL, CAL, ST and AEC contributed to data collection and processing. CMG, TT, and RW contributed to data analysis and presentation of results in figures and tables. RCW, JLS, SSK, and MPP assisted in study design. MPP acquired funding for the larger center grant that funded the project. All authors edited the manuscript and provided feedback on the original draft.

## Data availability

All data used in the analyses described in this paper are available in **Supplementary Materials**. **Code availability**

Model fitting code is publicly available in Zajkowski, Kossut (47) and was used here with minimal modification.

## Ethics approval and consent to participate

This study was carried out in accordance with the Declaration of Helsinki and approved by the ethics committee of the WCG Institutional Review Board (#20211403). All participants gave written informed consent.

## Conflict of interest or competing financial interests

The authors have no competing interests to disclose.

## Data Availability

All data produced in the present study are available upon reasonable request to the authors.

## Supplementary Materials

### Supplementary Methods

**Detailed information about medications used to determine eligibility:**

Psychiatric medications permitted with a stable dosage/frequency for 6 or more weeks:

- SSRIs/SNRIs (e.g., Zoloft, Celexa, Pristiq, Trintellix)
- Tricyclic and other antidepressants (e.g., Viibryd)
- Beta-blockers (Propranolol)
- Anxiolytics (Buspar)
- Lamictal, Gabapentin, or other anticonvulsants if prescribed expressly for depression/anxiety

Medications requiring 48 hours of abstinence before study visit:

- Cannabis
- Benzodiazepines (e.g., Xanax, Clonopin, Ativan)
- Alcohol
- Antihistamines used for short-term anxiolytic properties as needed (e.g., Hydroxyzine)
- Seroquel if used for short-term anxiolytic properties as needed

Medications not permitted:

- Antipsychotics (e.g., Geodon, Abilify, Vraylar), unless stable dosage was determined to be below threshold for psychiatric treatment (e.g., 100mg of Seroquel nightly for insomnia)
- Anticonvulsants (e.g., Gabapentin, Topiramate) prescribed for purposes other than antidepressant or anxiolytic treatment
- Stimulants (e.g., Adderall, Armodafinil, Modafinil, Vyvanse); Phentermine (amphetamine-like appetite suppressant)

**Table S1.**
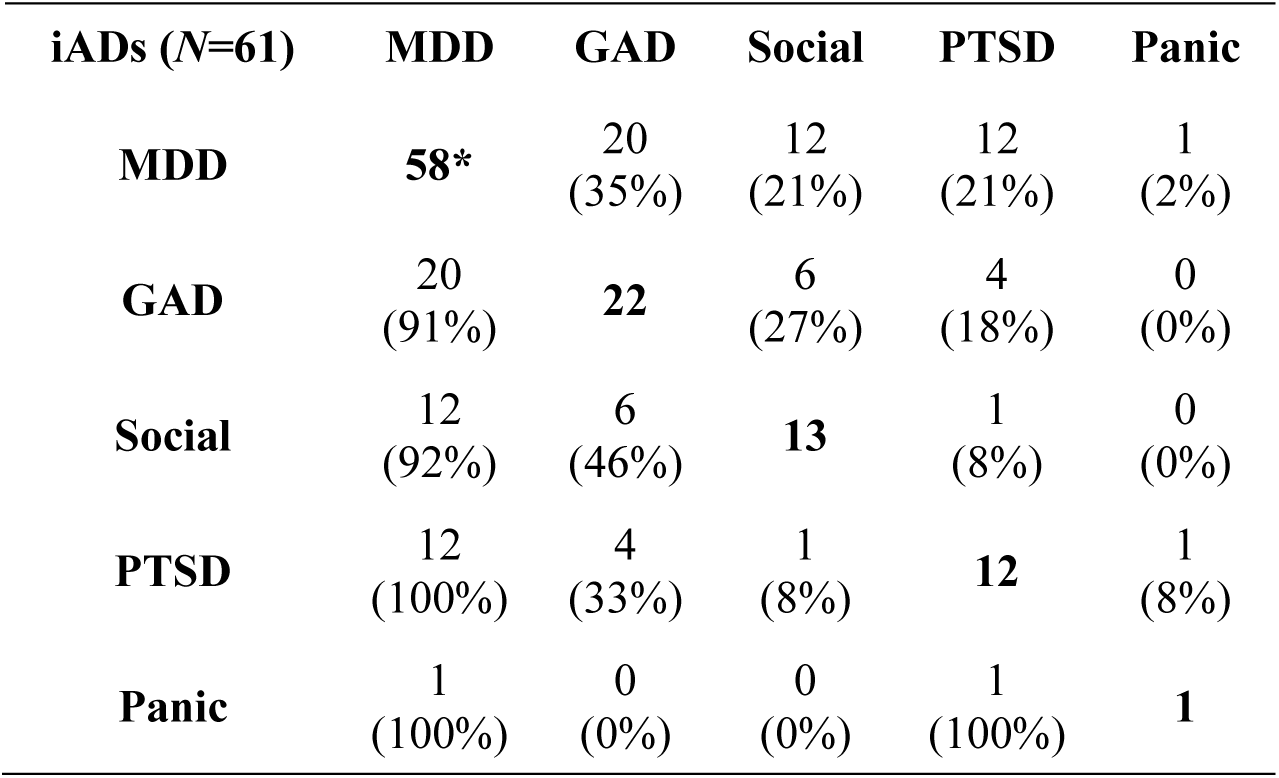
Number and proportion of individuals meeting criteria for specific affective disorders in the high-anxiety group.

| **iADs (N=61)** | **MDD** | **GAD** | **Social** | **PTSD** | **Panic** |
| --- | --- | --- | --- | --- | --- |
| **MDD** | **58\*** | 20<br>(35%) | 12<br>(21%) | 12<br>(21%) | 1<br>(2%) |
| **GAD** | 20<br>(91%) | **22** | 6<br>(27%) | 4<br>(18%) | 0<br>(0%) |
| **Social** | 12<br>(92%) | 6<br>(46%) | **13** | 1<br>(8%) | 0<br>(0%) |
| **PTSD** | 12<br>(100%) | 4<br>(33%) | 1<br>(8%) | **12** | 1<br>(8%) |
| **Panic** | 1<br>(100%) | 0<br>(0%) | 0<br>(0%) | 1<br>(100%) | **1** |

Note. MINI diagnoses were given up to 12 months prior to study participation. Participants in this group were required to have a current or lifetime diagnosis of one or more anxiety/depressive disorder(s) and OASIS > 8 for eligibility. Numbers on the diagonal indicate the total number of participants with lifetime occurrence of a given diagnosis. Numbers below the diagonal indicate participants with a given comorbidity. Percentages reflect the proportion of participants with a given comorbidity out of the total number of individuals with the diagnosis labelled on the respective rows. For example, values within the cell corresponding to row=GAD and column=MDD indicates that 91% of individuals with GAD also had MDD; whereas the entry for row=MDD and column=GAD indicates that 35% of individuals with MDD also had GAD. Thus, the counts are redundant above and below the diagonal, while the percentages are not. MDD=major depressive disorder, GAD=generalized anxiety disorder, Social=social anxiety disorder, PTSD=post-traumatic stress disorder, Panic=panic disorder.

*Of those with a lifetime diagnosis of MDD, 20 had current MDD, 30 were in partial remission, and 8 were in full remission.

**Table S2.** Sample Demographics.

| **Demographics** | **Healthy Comparisons**<br>**(N=58)** | **Affective Disorders**<br>**(N=61)** | **Sample**<br>**Comparison** |
| --- | --- | --- | --- |
| Age (mean (SD)) | 35.41 (13.08) | 33.44 (10.97) | $t(117) = 0.89, p = 0.374$ |
| Gender Identity = Woman | 42 (72.4%) | 46 (75.4%) | $\chi^2(1) = 0.03, p = .870$ |
| Sexual Orientation (counts) | | | $\chi^2(3) = 4.07, p = 0.254$ |
| Bisexual | 3 (5.2%) | 5 (8.5%) |  |
| Lesbian | 0 (0.0%) | 3 (5.1%) |  |
| None of these describe me | 1 (1.7%) | 2 (3.4%) |  |
| Straight, that is, not gay or lesbian, etc. | 54 (93.1%) | 49 (83.1%) |  |
| Education | | | $\chi^2(7) = 4.47, p = 0.724$ |
| GED or equivalent | 0 (0.0%) | 1 (1.6%) |  |
| High school graduate | 1 (1.7%) | 4 (6.6%) |  |
| Some college, no degree | 6 (10.3%) | 8 (13.1%) |  |
| Associate's degree: occupational, technical, or vocational program | 4 (6.8%) | 5 (8.2%) |  |
| Associate's degree: academic program | 3 (5.2%) | 4 (6.6%) |  |
| Bachelor's degree | 25 (43.1%) | 18 (29.5%) |  |
| Master's degree | 7 (12.1%) | 7 (11.5%) |  |
| Doctoral degree | 1 (1.7%) | 1 (1.6%) |  |
| Information Unavailable | 11 (19.0%) | 13 (21.3%) |  |
| Ethnicity = Not Hispanic/Latino | 52 (89.7%) | 54 (88.5%) | $\chi^2(1) = 0, p = 1$ |
| Race | | | $\chi^2(5) = 1.327, p = 0.932$ |
| American Indian or Alaska Native | 8 (13.8%) | 8 (13.6%) |  |
| Asian | 4 (6.9%) | 3 (5.1%) |  |
| Black or African American | 3 (5.2%) | 4 (6.8%) |  |
| Native Hawaiian or Pacific Islander | 1 (1.7%) | 0 (0.0%) |  |
| Other | 3 (5.2%) | 3 (5.1%) |  |
| White | 39 (67.2%) | 41 (69.5%) |  |

### Participant Withdrawals

The sensitivity assessment served not only as a way to obtain individual anxiety ratings to the breathing resistances, but also to allow initial exposure to the mask before participants began the Horizon Task. Seven participants who consented for the study withdrew either during or after the sensitivity assessment because of discomfort with the breathing mask. Of the 7 individuals who consented for this project and withdrew before completion due to tolerability of the breathing resistance, 5 were iADs and 2 were HCs. The iADs who withdrew were numerically older (M=44, SD=14.09) than those who did not and had comparable OASIS scores (M=8.5, SD=1.29) and numerically lower PHQ scores (M=8.25, SD=4.79). All 5 were diagnosed with MDD, 4 GAD, 1 PTSD, and 1 social anxiety disorder. Two were taking psychiatric medications. All 7 participants who withdrew were female. The small number of individuals who withdrew did not allow for formal statistical comparison to those who remained in the study.

### Measure Details

#### State-Trait Anxiety Inventory (STAI-State/Trait)

The STAI consists of two sets of 20 items designed to assess state and trait anxiety, respectively. Respondents were asked to rate each item on a scale of 1 (“not at all” for state anxiety; “almost never” for trait anxiety) to 4 (“very much so” for state anxiety; “almost always” for trait anxiety) based on how much they agree or disagree with the associated statement. The STAI has shown good to excellent internal consistency (Cronbach’s alphas ranging from .86 to .96; (1)).

#### Overall Anxiety Sensitivity and Impairment Scale (OASIS)

The OASIS is a 5-item self-report measure designed to assess anxiety-related impairment across various areas of life over the past week. Total scores are obtained by summing scores of individual items (0-4), ranging from 0 to 20, where higher scores indicate greater impairment. The OASIS has shown high internal consistency, with a Cronbach’s alpha of .80 (2).

#### Patient Health Questionnaire (PHQ-9)

The PHQ is a screening tool for depression and includes 9 self-report items, which correspond to the diagnostic criteria of major depressive disorder. Each item statement is rated on a 4-point Likert Scale, ranging from 0 (not at all) to 3 (nearly every day) to describe the extent to which a depressive symptom has bothered them in the past 2 weeks. The total score reflects level of depression severity. The PHQ-9 has been found to show good internal consistency (α = .89; (3)).

#### Childhood Trauma Questionnaire (CTQ)

The CTQ short form (4) includes 28 self-report items that measure childhood abuse and neglect through five subscales: physical abuse, emotional abuse, sexual abuse, physical neglect, and emotional neglect. Each item is scored using a Likert scale, ranging from 1 (“never true”) to 5 (“very often true”), with relevant items summed to provide each subscale score. The total score is the mean of the subscale scores.

Higher scores indicate higher levels of childhood abuse and neglect.

#### Cognitive Reflection Test (CRT-7)

The CRT-7 (5) is comprised of 7 short questions that are designed to assess trait cognitive reflectiveness. Lower reflectiveness is operationalized as an individual’s tendency to provide an immediate intuitive answer that is incorrect, when more detailed reflection on the content of the question would lead to a less intuitive but correct answer. Example item: “Jerry received both the 15^th^ highest and the 15^th^ lowest mark in the class. How many students are in the class?” (intuitive answer: 30; correct answer: 29). Scores are based on the number of correct answers. The CRT-7 has shown acceptable reliability (α=.72; (5)).

### Sensitivity Assessment

In addition to self-reported anxiety levels, participants also gave responses to a series of other related questions immediately after exposure to each breathing resistance during the sensitivity protocol (exact wording listed below). Self-Assessment Manikin items assessed happiness and excitement (6) rated on a 5-point scale, while the others were rated similarly to self-reported anxiety (11-point scale). See **Figure S2** below for associated bar plots demonstrating ratings across resistance levels.

*“How calm or excited did you feel during loaded breathing?”* (on a scale from 1 to 5)

*“How happy or unhappy did you feel during loaded breathing?”* (on a scale from 1 to 5)

*“How difficult did it feel to breathe?”* (0-no difficulty; 10-maximal difficulty you could tolerate)

*“How much fear did you feel while breathing?”* (0-no fear at all; 10-maximal fear you could tolerate)

*“How unpleasant did it feel to breathe?”* (0-not at all unpleasant; 10-maximal unpleasantness you could tolerate)

### Computation Model and Model Fitting

#### Learning Model

The learning model is based on the Kalman filter (7). Independently for each game, it describes how participants update their beliefs about the mean reward value for each option (bandit) after observing each forced-choice outcome. The model assumes that rewards (𝑟) are generated from a Gaussian distribution with a fixed standard deviation (𝜎_𝑟_) and a mean (𝑅) that differs between options and can change over time. Changes in the mean are determined by a Gaussian random walk with mean 0 and standard deviation (𝜎_𝑑_), reflecting how far the mean may have drifted from its previous value after each choice in a game. Importantly, the true value of 𝜎_𝑑_ in this task is 0.

Consistent with this, task instructions inform participants that the average reward for each option is stable within a game. Incorporating this parameter thus allows for the possibility of suboptimal learning as well as a recency bias in mean estimates.

For bandit 𝑖 on trial 𝑡, estimation of the mean reward (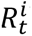) is updated based on a reward prediction error (i.e., the difference between the observed reward and the current mean estimate). This prediction error is scaled by a time-varying learning rate (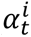), which changes as a function of one’s current uncertainty about the mean reward value for a given option, 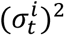, and the expected outcome variance or noise around the mean reward value (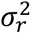), as follows:

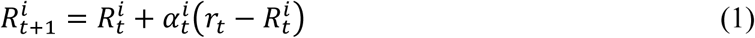

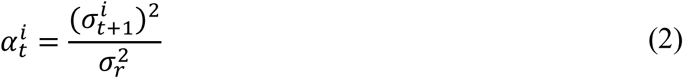

This entails that the learning rate will be slower if one expects the outcome noise around the mean (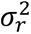) to be high, and that learning rate will decrease as one’s confidence in the mean value increases (i.e., as 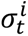 decreases with each new outcome that is observed).

Changes in the value for 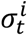 (and therefore 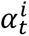) after each observation further depend on the expected variance or drift (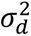) in the underlying mean:

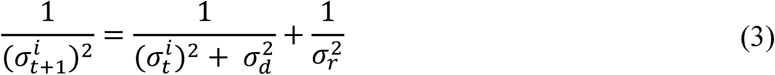

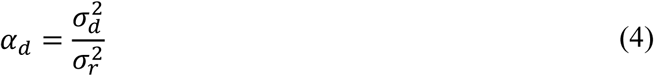

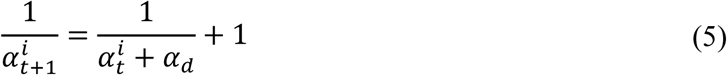

Equation 3 entails that uncertainty around the mean reward value will decrease when 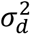 and 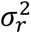 take on sufficiently small values, but that it will increase when they take on larger values.

Equations 4 and 5 represent this in terms of learning rates. Here, it can be seen that learning rates will remain relatively higher when individuals think the reward mean is likely to have changed (i.e., higher values of 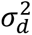). Specifically, learning rate depends on the ratio of the drift variance and the noise variance. Thus, learning rate will be higher if one expects that surprising outcomes are more likely due to changes in the mean than due to outcome noise.

If bandit 𝑖 is *not* played on trial 𝑡, the estimated mean is instead assumed to stay the same, while the uncertainty in that estimate grows in accordance with 𝜎_𝑑_, as this entails the mean may have changed:

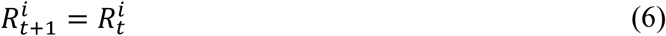

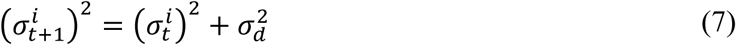

Overall, the Kalman filter model therefore has four free parameters: the initial expected reward mean (𝑅_0_), the initial expected uncertainty around that mean (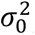), the expected drift variance (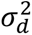), and the expected outcome noise (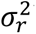). For ease of interpretability, and because only three of these four parameters can be estimated from behavioral data on this task, previous studies have chosen to reparametrize the three variance parameters in terms of two learning rates (8, 9). We follow this same convention here. The first parameter in this approach is an initial learning rate (𝛼_0_), which corresponds to a ratio of the three variances:

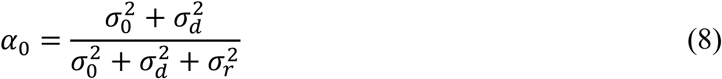

The second parameter is an asymptotic learning rate (𝛼_∞_), which is the steady state value of 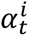 if bandit 𝑖 were chosen ad infinitum. This can be calculated in terms of 𝛼_𝑑_ as follows:

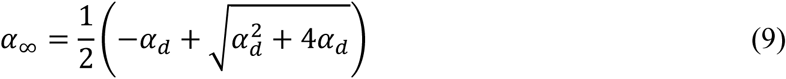

This reparameterization then constrains both learning rate values to between 0 and 1. In place of estimating the values of each of the variances, this approach estimates these two learning rates when fitting the model to behavioral data (i.e., where the model is implemented in terms of the learning rate update rule shown in equation 5). Here, 𝛼_0_ estimates can be seen to indirectly entail expected values for 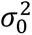 in relation to the other variances, while 𝛼_∞_ indirectly entails the expected ratio between 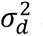 and 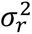. The value for 𝑅_0_ can either be estimated or fixed to a specific starting value. Because this parameter is known to trade off with other parameters in the decision model (see below) in explaining directed exploration, we here simply fix 𝑅_0_ to a neutral value of 50. As each of the 80 games is independent, and participants are informed of this, it is assumed that no learning carries over between games or task runs. Thus, 𝑅_0_ is reset to this value for each of the new bandits at the start of each game.

#### Decision Model

After estimating the mean reward (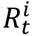) of each bandit from the forced-choice outcomes, the participant is assumed to choose between the two bandits according to a logistic choice rule:

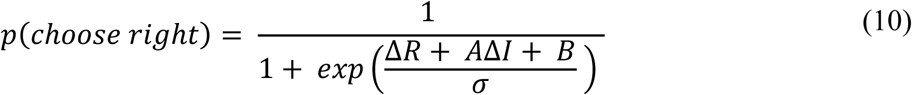

Here, Δ𝑅 is the difference in expected reward values between the two bandits. The Δ𝐼 term instead denotes the difference in available information between the two bandits (i.e., where choosing the bandit with only one observed forced-choice outcome would be more informative than choosing the bandit with three forced-choice outcomes). In this case, Δ𝐼 is assigned a value of 1 when the left bandit is more informative, -1 when the right bandit is more informative, and 0 for equal information games. This choice rule also includes three parameters. The information bonus parameter (*A*) moderates how strongly the difference in available information (Δ𝐼) influences choice (i.e., greater values promote selection of the bandit where only one forced-choice outcome has been observed). The decision noise parameter (𝜎) controls the probability that choices with lower expected values will be chosen (i.e., higher values lead to less value-sensitive [more random] choices). The spatial bias parameter (*B*) estimates potential biases toward simply choosing the option on one side vs. the other. Separate values of each parameter are estimated for each horizon condition (i.e., H1 and H6). The spatial bias and decision noise parameters are further separated by uncertainty condition (i.e., separate estimates for equal and unequal information games). The information bonus parameter can only be estimated for unequal information games. Directed exploration (DE) is then taken to be the increase in information bonus values from H1 (when information is not useful in guiding future choice) to H6 (when information is useful in guiding future choices). That is, 𝐷𝐸 = 𝐴_𝐻6_ − 𝐴_𝐻1_. By the same reasoning, random exploration (RE) is defined as the increase in decision noise when information becomes useful in guiding future choice beyond an individual’s baseline level of decision noise in H1. That is, 𝑅𝐸 = 𝜎_𝐻6_ − 𝜎_𝐻1_. Note that, while decision noise is estimated in both equal and unequal information conditions, we restrict analyses of decision noise and random exploration to estimates for the equal information condition to minimize potentially confounding interactions with directed exploration behavior.

#### Model Fitting

A previously used hierarchical Bayesian model was used to fit each parameter to the behavior of each participant (8, 9). In this approach, it is assumed that each parameter for each participant is taken from the same second-level prior distribution, and is estimated using a Markov Chain Monte Carlo (MCMC) sampling procedure. Second-level priors are in turn parameterized by hyperpriors set to have broad, minimally informative distributions (for specific values, see **Table S3**). This hierarchical model fitting approach used data from all participants across both resistance conditions to estimate parameters simultaneously (i.e., with single second-level priors). This choice was made to prevent potential inflation of group or condition differences that could occur if separate prior group or condition means were assumed.

Model fitting was performed using the JAGS package (Plummer, 2003) to run Markov Chain Monte Carlo simulations in MATLAB (psiexp.ss.uci.edu/research/programs_data/ jags/). This package draws samples from the posterior distribution given the observed behavioral data to approximate the posterior distribution over model parameters. Specifically, 1000 samples were generated from the posterior distribution over model parameters for each of 4 independent Markov chains (i.e., 4000 samples in total). We discarded the first 500 samples of each chain (i.e., burn-in = 500) that might impact the posterior distribution and generated posterior samples at a thin rate of 1.

**Table S3.** Computational Model Parameter Prior Values.

| Parameter | Prior | Hyperparameters | Hyperpriors |
| --- | --- | --- | --- |
| Initial Learning Rate $\alpha_0$ | $\alpha_0 \sim \text{Beta}(a_{\alpha_0}, b_{\alpha_0})$ | $\theta_{\alpha_0} = \text{Beta}(a_{\alpha_0}, b_{\alpha_0})$ | $a_{\alpha_0} \sim \text{Uniform}(0.1, 10)$<br>$b_{\alpha_0} \sim \text{Uniform}(0.5, 10)$ |
| Asymptotic Learning Rate $\alpha_\infty$ | $\alpha_\infty \sim \text{Beta}(a_{\alpha_\infty}, b_{\alpha_\infty})$ | $\theta_{\alpha_\infty} = \text{Beta}(a_{\alpha_\infty}, b_{\alpha_\infty})$ | $a_{\alpha_\infty} \sim \text{Uniform}(0.1, 10)$<br>$b_{\alpha_\infty} \sim \text{Uniform}(0.5, 10)$ |
| Information Bonus $A$ | $A \sim \text{Gaussian}(\mu_A, \sigma_A)$ | $\theta_A = \text{Gaussian}(\mu_A, \sigma_A)$ | $\mu_A \sim \text{Gaussian}(0, 100)$<br>$\sigma_A \sim \text{Gamma}(1, 0.001)$ |
| Spatial Bias $B$ | $B \sim \text{Gaussian}(\mu_B, \sigma_B)$ | $\theta_B = \text{Gaussian}(\mu_B, \sigma_B)$ | $\mu_B \sim \text{Gaussian}(0, 100)$<br>$\sigma_B \sim \text{Gamma}(1, 0.001)$ |
| Decision Noise $\sigma$ | $\sigma \sim \text{Gamma}(k_\sigma, \lambda_\sigma)$ | $\theta_\sigma = \text{Gamma}(k_\sigma, \lambda_\sigma)$ | $k_\sigma \sim \text{Exp}(0.1)$<br>$\lambda_\sigma \sim \text{Exp}(10)$ |

## Supplementary Results

### Model Validation

Confirmatory correlations between model-free and model-based metrics of behavior indicated expected relationships. In particular, IB1 and IB6 were positively associated with the probability of choosing the high information option in H1 and H6 games, respectively (*rs*>.86, *ps*<.001 for both resistance conditions). IB1 was also positively associated with the probability of the choosing the low mean bandit in both resistance conditions (*rs*>.33, *ps*<.001) while IB6 was only associated with the probability of choosing the low mean bandit for H6 in the resistance condition (*r*=.19, *p*=.034). As expected, DN parameters were positively associated with the probability of choosing the low mean bandit in their respective game types (i.e., H1, H6; *rs*>.29, *ps*<.002) and were largely not associated with the probability of the choosing the high information side on unequal information games, apart from DN for H1 unequal games in the no-resistance condition (*r*=-.33, *p*<.001). Finally, learning rates were negatively correlated with the probability of choosing the low mean bandit (*rs*<-.43, *ps*<.001) and positively associated with points won in each trial condition (*rs*>.45, *ps*<.001). Most of the parameters across both resistance conditions were not associated with reaction times (RTs; *ps*>.05), except for 𝜶_𝟎_in the no-resistance condition (*r*=.19, *p*=.038).

### Breathing Resistance Sensitivity

Anxiety ratings during the breathing resistance sensitivity protocol are shown in **Figure S1**. In the LME of anxiety including resistance level, group, and their interaction as regressors, all effects were significant (*Fs*>5.77, *ps*<.001). Post-hoc contrasts indicated that 1) iADs had higher self-reported anxiety levels than HCs at each resistance level (*ps*<.013), 2) that anxiety at resistance levels 40, 60, and 80 were not significantly different from each other in HCs (*ps*>.076), and 3) that each increase in resistance level resulted in a significant increase in anxiety for the iADs, but anxiety in HCs did not change from 40 through 80. These results indicate that anxiety induction was successful across participants, and that iADs showed greater sensitivity than HCs.

LMEs for self-reported anxiety scores at baseline and during both task runs (**Figure 2**) showed significant effects of group (*F*(1,117)=49.44, *p*<.001), resistance condition (*F*(2,234)=105.89, *p*<.001), and their interaction (*F*(2,234)=8.04, *p*<.001). Post-hoc analysis revealed that: 1) iADs (estimated marginal mean [EMM]=3.06) reported higher anxiety ratings than HCs (EMM=1.03; *t*(117)=7.03, *p*<.001); 2), anxiety ratings were highest during the task condition with the added resistance (EMM=3.20), followed by the task condition without the resistance (EMM=1.63) and baseline ratings (EMM=1.31; *ps*<.022); and 3) self-reported anxiety ratings did not increase from baseline to the no-resistance task run for HCs (EMM_baseline_=0.53, EMM_no-resistance_=0.69; *p*=.440), but did for iADs (EMM_baseline_=2.08, EMM_no-resistance_=2.57; *p*=.013).

Similar results were found for complementary STAI-State ratings. Here there were main effects of group (*F*(1,117)=54.93, *p*<.001) and resistance (*F*(2,234)=85.51, *p*<.001), but no interaction (*F*(2,234)=0.56, *p*=.570). Post-hoc contrasts indicated that iADs reported higher anxiety (EMM=44.9) than HCs (EMM=33.1; *t*(117)=7.41, *p*<.001) and that anxiety during the run with the added breathing resistance (EMM=44.3) was higher than anxiety at baseline (EMM=35.7) and when no resistance was added (*ps*<.001), but baseline was not significantly different from the no resistance condition (*t*(234)=1.73, *p*=.085).

### Disentangling Relationships between Additional Model Parameters and Depression/Anxiety

Within HCs, linear regressions with RE as the outcome variable revealed no significant effects for STAI in either resistance condition. In iADs, there was a marginal effect of PHQ in the no-resistance condition with and without covariates (b=-0.13, *p*=.065 and b=-0.13, *p*=.071, respectively). We confirmed VIFs<4 for all predictors in this model. Confirmatory ridge regressions (minimizing potential multicollinearity and variance inflation effects) also showed relationships in the same directions as in the original linear model for both symptom measures, although here these were nonsignificant (i.e., noting the reduced statistical power afforded by this analysis approach).

Within HCs, a logistic regression for 𝜶_𝟎_ indicated a marginal negative association with STAI Trait in the resistance condition (z=-1.94, *p*=.053) without covariates, which became significant after adding covariates (z=-2.01, *p*=.045). No other significant effects were found for either resistance condition. In iADs, no significant effects were observed. Complementary nonparametric tests also revealed no additional associations in any case.

With respect to 𝜶_∞_, there were significant main effects of both PHQ (*Fs*>4.50, *ps*<.039) and STAI Trait (*Fs*>4.32, *ps*<.043) in HCs in both resistance conditions after adding covariates.

These relationships indicated a positive effect of STAI Trait (b=0.01) and a negative effect of PHQ (b=-0.02) on learning rates in the no-resistance condition, but these directions flipped during the resistance condition (b=-0.01 and b=0.03, respectively). No associations were found for iADs in either condition.

**Table S4.** Descriptive Statistics for Model Parameters (mean and SD)

|  | No resistance condition |  | Resistance condition |  |
| --- | --- | --- | --- | --- |
|  | HC | iADs | HC | iADs |
| DE | 5.27 (3.8) | 5.24 (3.68) | 4.28 (3.8) | 5.34 (3.44) |
| IB1 | -1.75 (0.95) | -1.64 (1.21) | -2.03 (1.14) | -1.57 (1.01) |
| IB6 | 3.51 (4.1) | 3.59 (3.89) | 2.25 (3.97) | 3.77 (3.7) |
| RE (equal information) | 1.91 (2.41) | 1.58 (1.89) | 1.52 (1.86) | 1.61 (1.81) |
| DN1 (equal information) | 2.05 (0.66) | 2.24 (0.74) | 2.18 (0.69) | 2.32 (0.75) |
| DN6 (equal information) | 3.96 (2.19) | 3.81 (1.69) | 3.7 (1.81) | 3.93 (1.63) |
| RE (unequal information) | 2.8 (2.72) | 2.96 (2.97) | 2.22 (2.6) | 2.79 (2.31) |
| DN1 (unequal information) | 2.74 (1.46) | 2.69 (1.33) | 3.04 (1.45) | 2.9 (1.61) |
| DN6 (unequal information) | 5.54 (2.63) | 5.65 (2.62) | 5.26 (2.31) | 5.69 (2.24) |
| $\alpha_0$ | 0.52 (0.28) | 0.46 (0.31) | 0.53 (0.28) | 0.41 (0.31) |
| $\alpha_\infty$ | 0.33 (0.12) | 0.28 (0.13) | 0.34 (0.13) | 0.27 (0.12) |

Note: DE=directed exploration, IB1/IB6=information bonus for H1/H6 conditions, RE=random exploration, DN1/DN6=decision noise in H1/H6 conditions, $\alpha_0$ =initial learning rate, $\alpha_\infty$ =asymptotic learning rate. As these are not used in subsequent statistical analyses, DN1/DN6 values for the unequal information condition are reported in **Supplementary Materials (Figure S3)**. \* $p<.05$ , \*\* $p<.01$ , \*\*\* $p<.001$ .

**Table S5.**
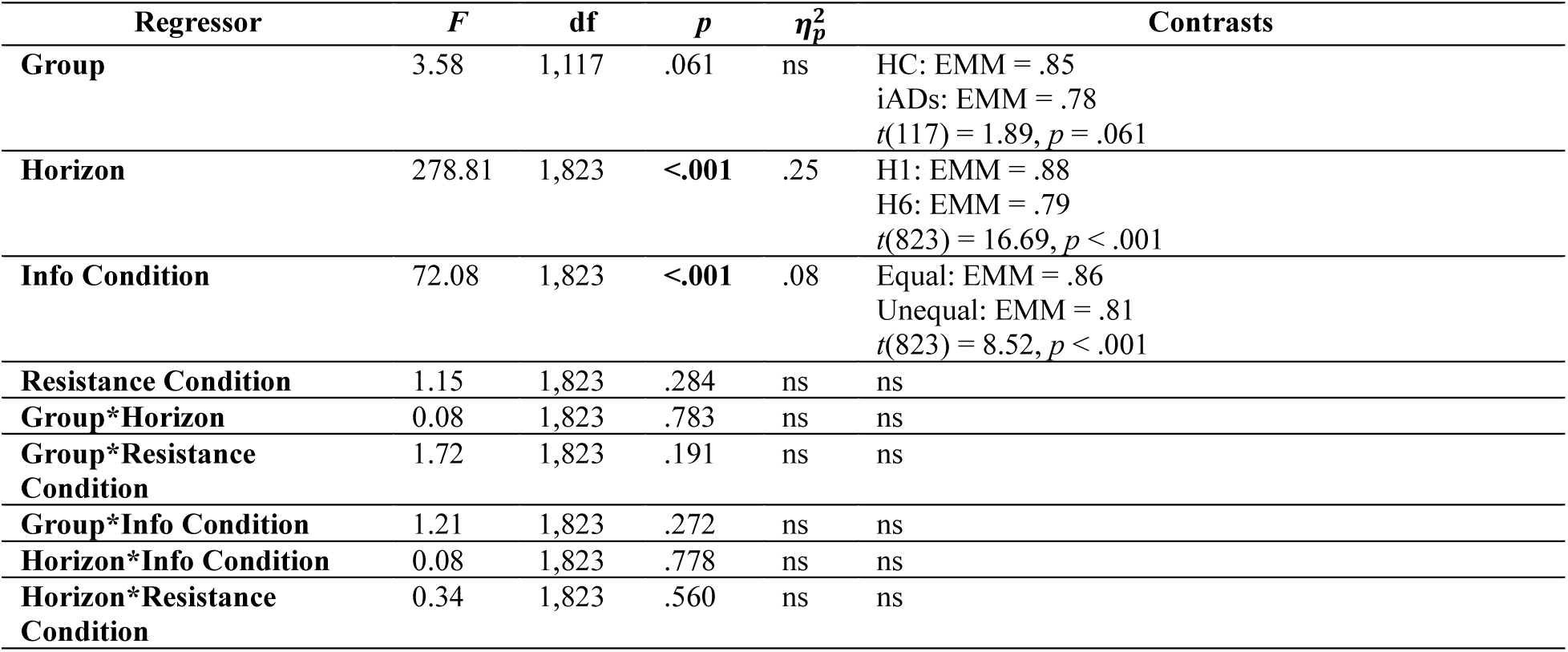
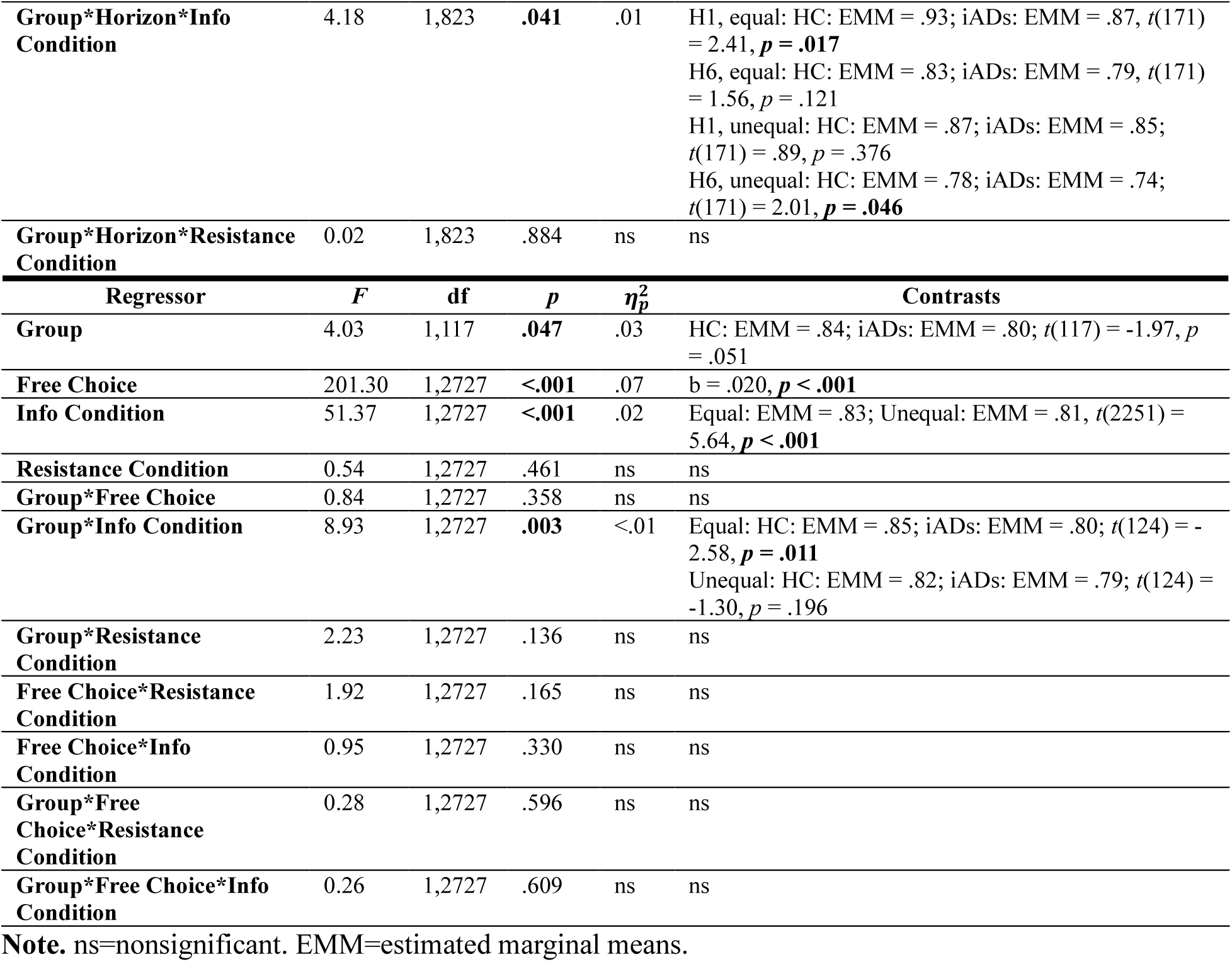
Statistics for models of accuracy on the first free choice (top) and accuracy across all free choices in the Horizon 6 condition (bottom).

| Regressor | *F* | df | *p* | $\eta_p^2$ | Contrasts |
| --- | --- | --- | --- | --- | --- |
| Group | 3.58 | 1,117 | .061 | ns | HC: EMM = .85<br>iADs: EMM = .78<br>$t(117) = 1.89, p = .061$ |
| Horizon | 278.81 | 1,823 | <.001 | .25 | H1: EMM = .88<br>H6: EMM = .79<br>$t(823) = 16.69, p < .001$ |
| Info Condition | 72.08 | 1,823 | <.001 | .08 | Equal: EMM = .86<br>Unequal: EMM = .81<br>$t(823) = 8.52, p < .001$ |
| Resistance Condition | 1.15 | 1,823 | .284 | ns | ns |
| Group*Horizon | 0.08 | 1,823 | .783 | ns | ns |
| Group*Resistance Condition | 1.72 | 1,823 | .191 | ns | ns |
| Group*Info Condition | 1.21 | 1,823 | .272 | ns | ns |
| Horizon*Info Condition | 0.08 | 1,823 | .778 | ns | ns |
| Horizon*Resistance Condition | 0.34 | 1,823 | .560 | ns | ns |

| **Group\*Horizon\*Info Condition** | 4.18 | 1,823 | **.041** | .01 | H1, equal: HC: EMM = .93; iADs: EMM = .87, $t(171) = 2.41, p = .017$<br>H6, equal: HC: EMM = .83; iADs: EMM = .79, $t(171) = 1.56, p = .121$<br>H1, unequal: HC: EMM = .87; iADs: EMM = .85; $t(171) = .89, p = .376$<br>H6, unequal: HC: EMM = .78; iADs: EMM = .74; $t(171) = 2.01, p = .046$ |
| --- | --- | --- | --- | --- | --- |
| **Group\*Horizon\*Resistance Condition** | 0.02 | 1,823 | .884 | ns | ns |
| **Regressor** | **F** | **df** | **p** | **$\eta_p^2$** | **Contrasts** |
| **Group** | 4.03 | 1,117 | **.047** | .03 | HC: EMM = .84; iADs: EMM = .80; $t(117) = -1.97, p = .051$ |
| **Free Choice** | 201.30 | 1,2727 | **<.001** | .07 | b = .020, **$p < .001$** |
| **Info Condition** | 51.37 | 1,2727 | **<.001** | .02 | Equal: EMM = .83; Unequal: EMM = .81, $t(2251) = 5.64, p < .001$ |
| **Resistance Condition** | 0.54 | 1,2727 | .461 | ns | ns |
| **Group\*Free Choice** | 0.84 | 1,2727 | .358 | ns | ns |
| **Group\*Info Condition** | 8.93 | 1,2727 | **.003** | <.01 | Equal: HC: EMM = .85; iADs: EMM = .80; $t(124) = -2.58, p = .011$<br>Unequal: HC: EMM = .82; iADs: EMM = .79; $t(124) = -1.30, p = .196$ |
| **Group\*Resistance Condition** | 2.23 | 1,2727 | .136 | ns | ns |
| **Free Choice\*Resistance Condition** | 1.92 | 1,2727 | .165 | ns | ns |
| **Free Choice\*Info Condition** | 0.95 | 1,2727 | .330 | ns | ns |
| **Group\*Free Choice\*Resistance Condition** | 0.28 | 1,2727 | .596 | ns | ns |
| **Group\*Free Choice\*Info Condition** | 0.26 | 1,2727 | .609 | ns | ns |

**Note.** ns=nonsignificant. EMM=estimated marginal means.

## Supplementary Figures

**Figure S1.**
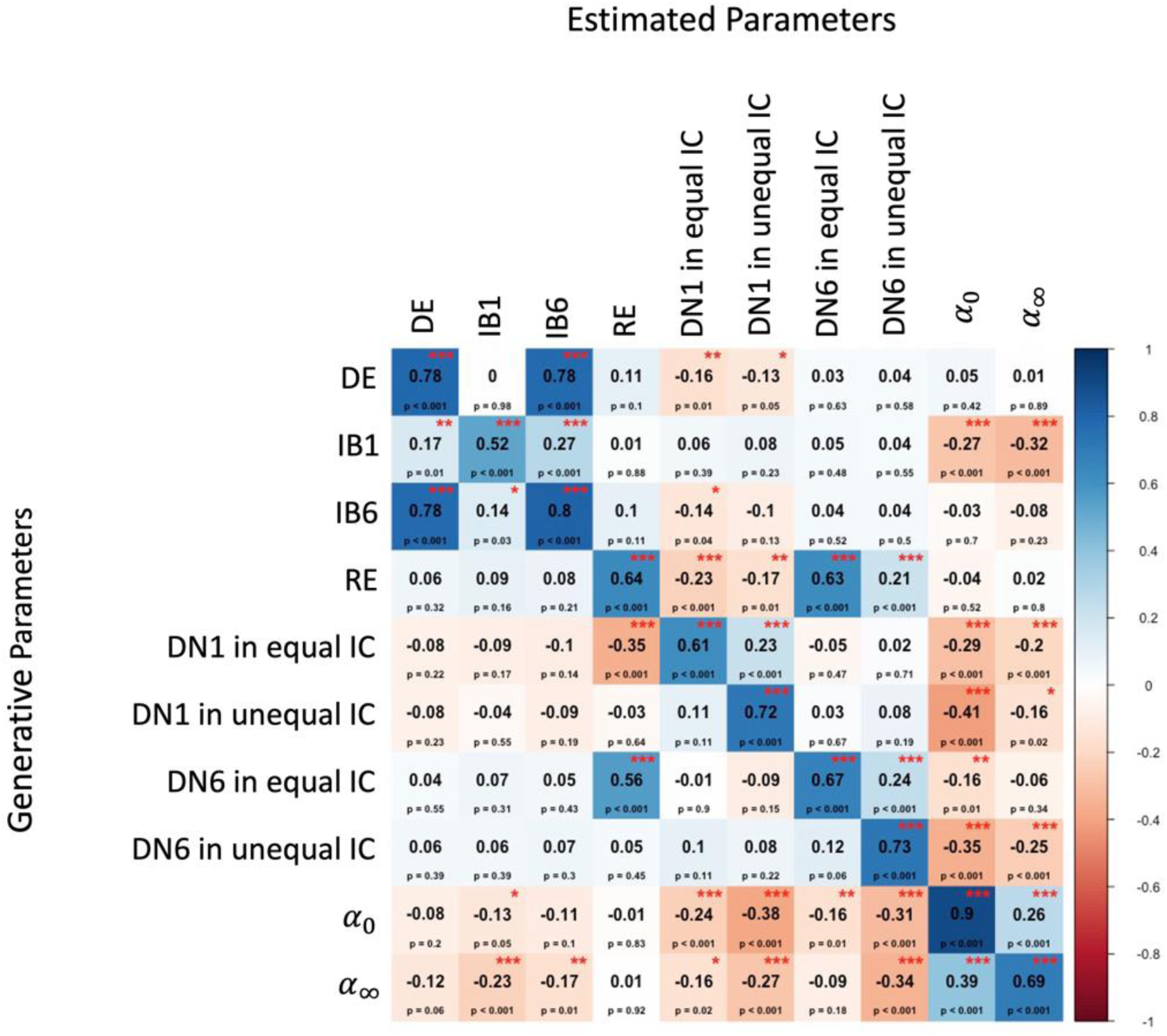
Parameter recoverability analysis indicating correlations between estimated (fit) and generated (simulated) parameter values and associated significance levels. DE=directed exploration, IB1/IB6=information bonus for H1/H6 conditions, RE=random exploration, DN1/DN6=decision noise in H1/H6 conditions, IC=information condition, 𝛼_0_=initial learning rate, 𝛼_∞_=asymptotic learning rate. \**p*<.05, \*\**p*<.01, \*\*\**p*<.001.

**Figure S2.**
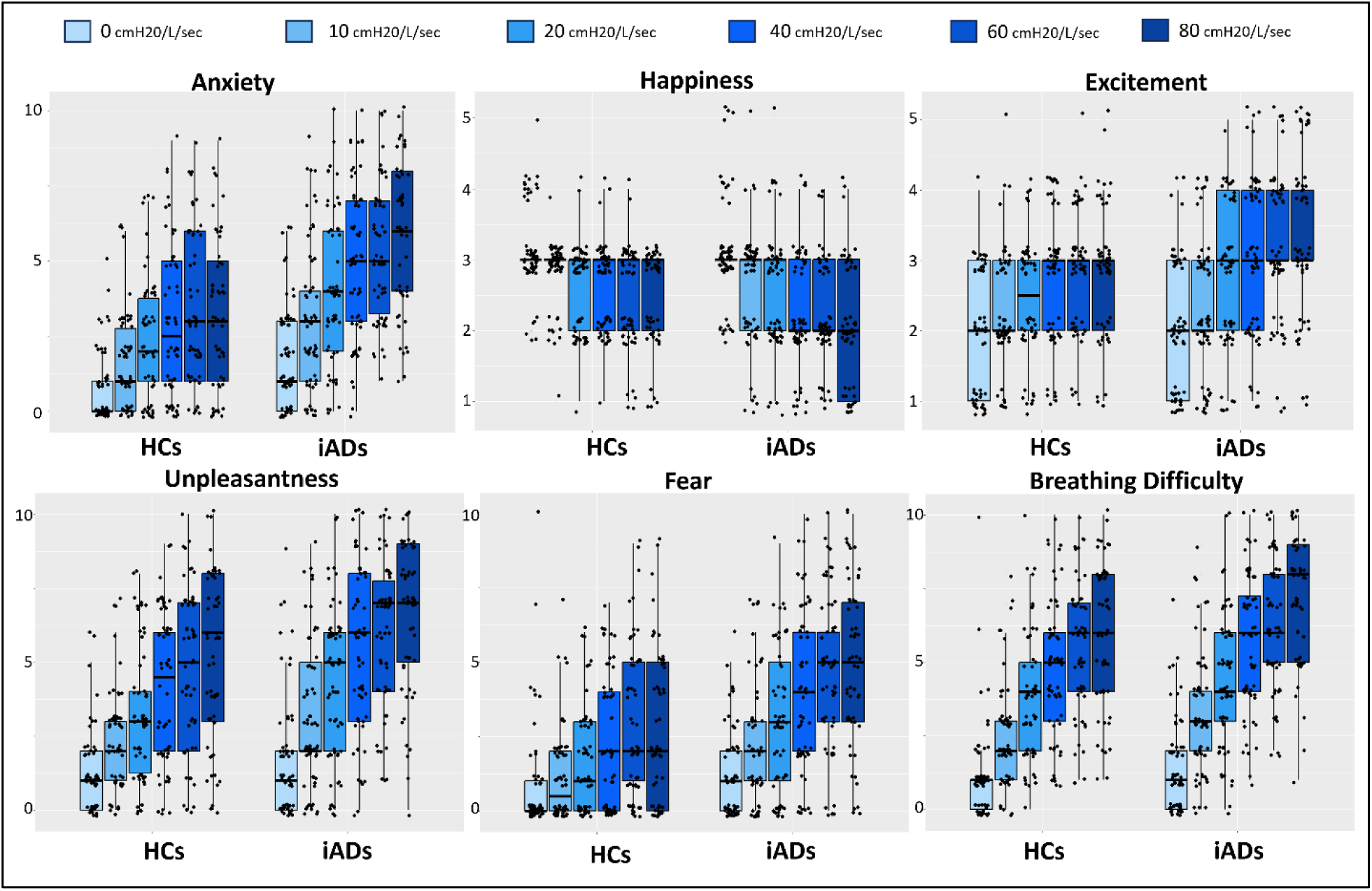
Participant ratings during the breathing resistance sensitivity protocol across 6 levels of breathing resistance (0 to 80 cmH2O/L/sec). Boxplots show medians and quartiles as well as individual datapoints.

**Figure S3.**
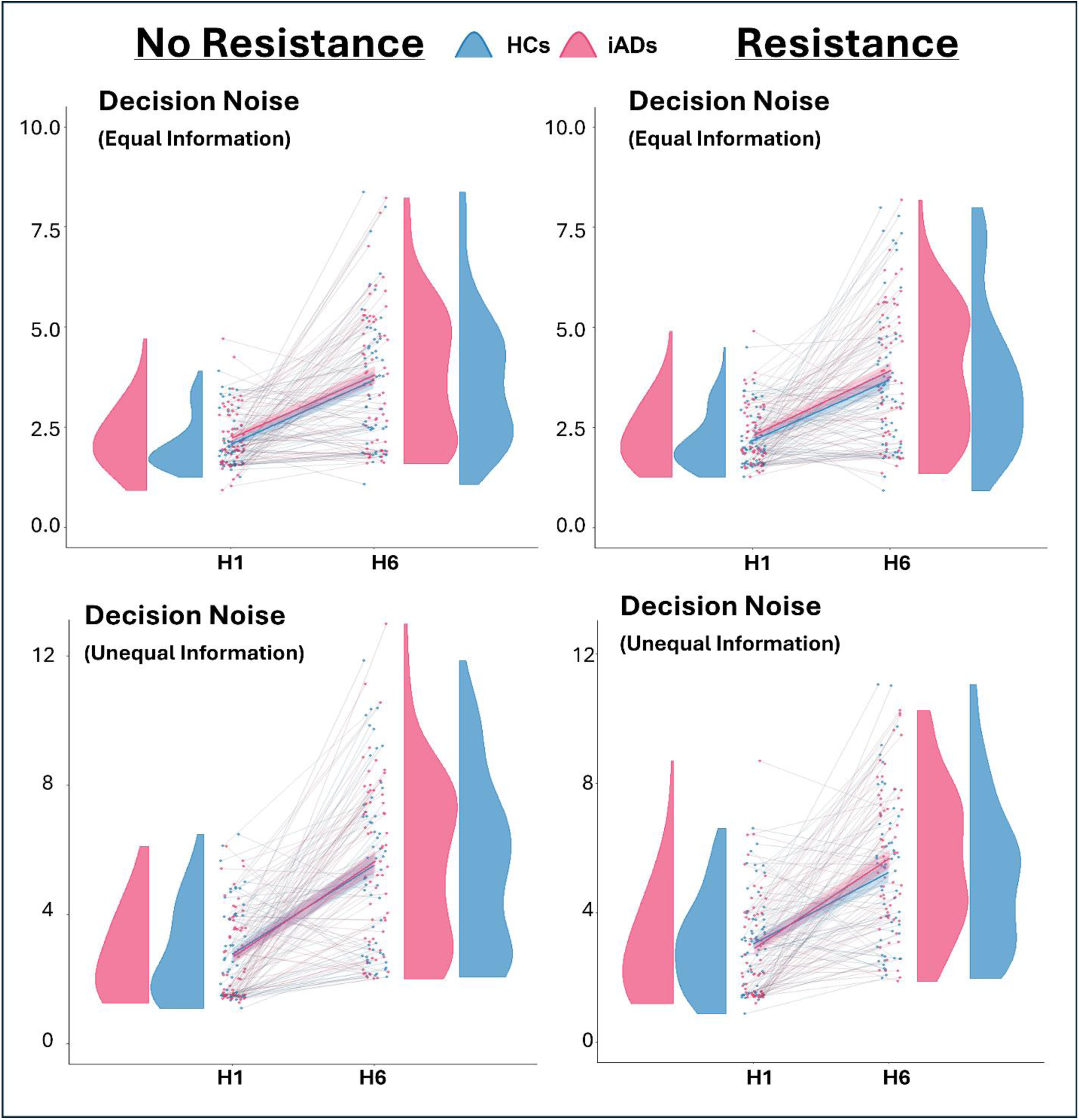
Decision noise parameters separated by horizon, group, and resistance condition.

**Figure S4.**
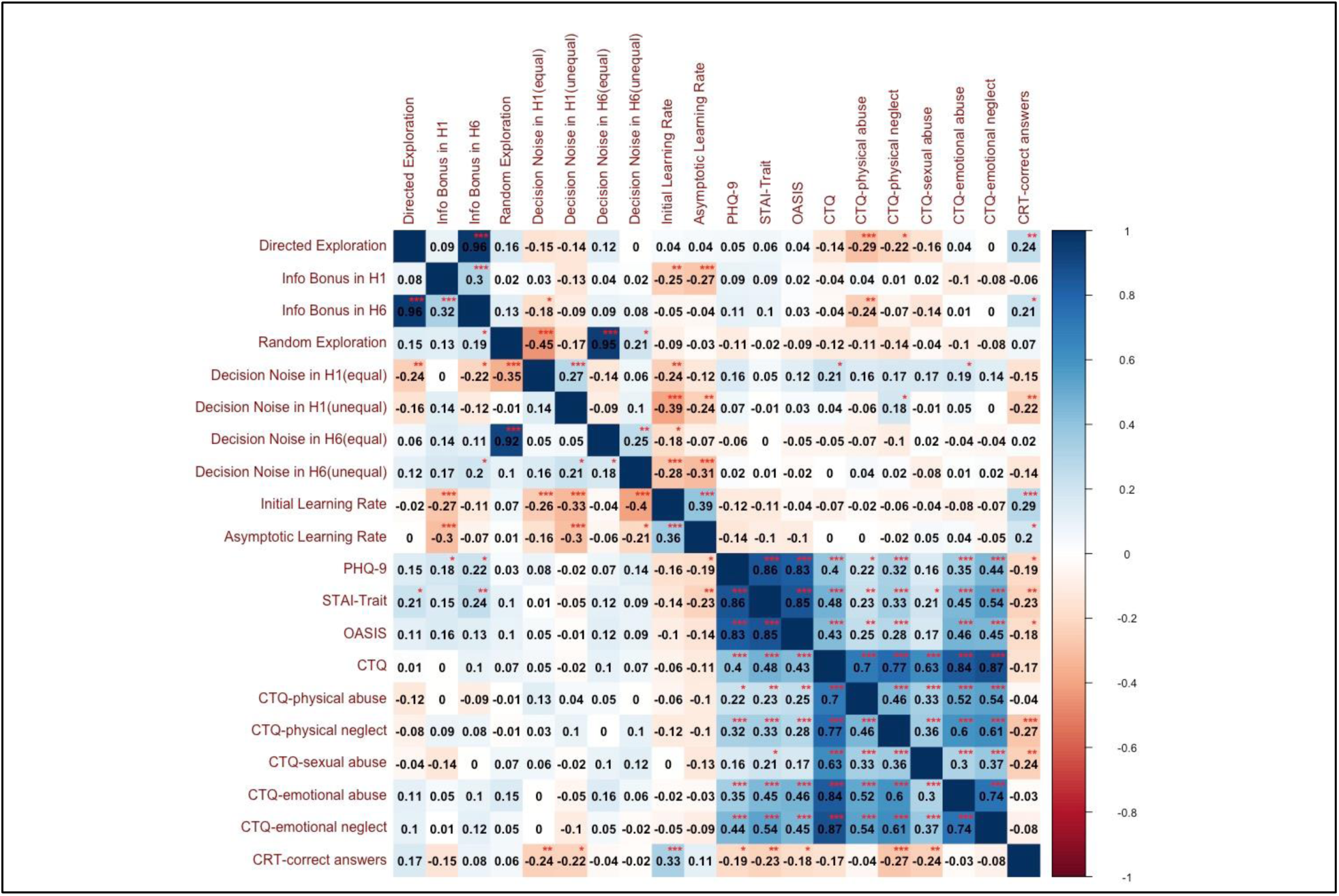
Post-hoc correlations between model parameters and self-report measures. Statistics below the diagonal indicate correlations with parameters in the resistance condition, while those above the diagonal are from the no-resistance condition. Note that correlations between 𝛼_0_ and other variables were conducted using Spearman correlations due to non-normality.

**Figure S5.**
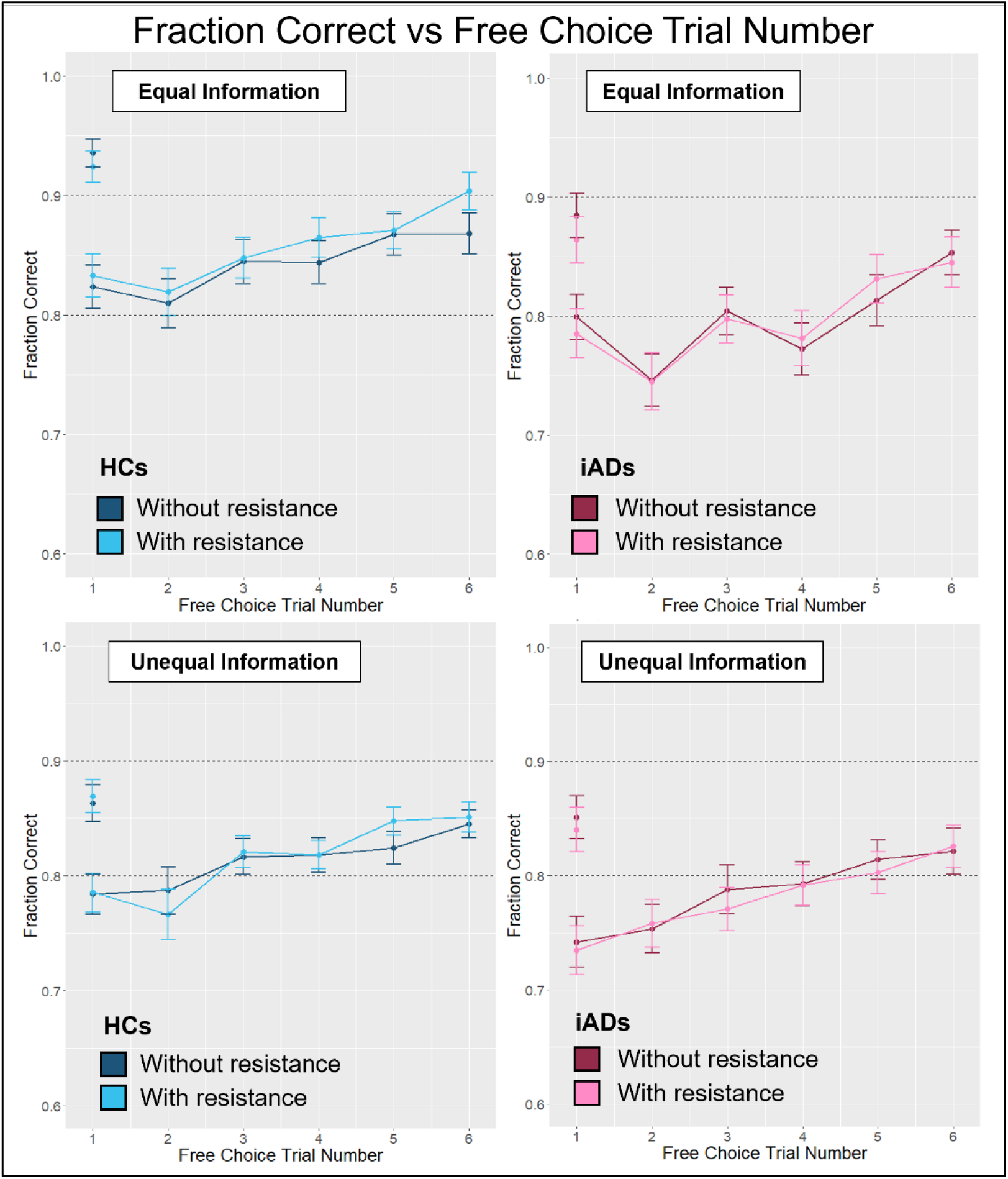
Fraction of correct responses (i.e., option with higher generative mean) for each free choice in H1 and H6 trials. Color shades indicate task performance with (light) and without (dark) breathing resistance separately in HCs (left/blue) and iADs (right/pink). Plots are also shown separately for equal (top) and unequal (bottom) information conditions.

## References

1. Barlow DH, Allen LB, Choate ML. Toward a unified treatment for emotional disorders - republished article. Behav Ther. 2016;47(6):838–53.

2. Hayes S, Strosahl K, Wilson K. Acceptance and commitment therapy: An experiential approach to behaviour change. 2003. New York: Guilford Press.

3. Sharp PB, Eldar E. Computational models of anxiety: nascent efforts and future directions. Current Directions in Psychological Science. 2019;28(2):170–6.

4. Lloyd A, Roiser JP, Skeen S, Freeman Z, Badalova A, Agunbiade A, et al. Reviewing explore/exploit decision-making as a transdiagnostic target for psychosis, depression, and anxiety. Cogn Affect Behav Neurosci. 2024.

5. Browning M, Behrens TE, Jocham G, O’Reilly JX, Bishop SJ. Anxious individuals have difficulty learning the causal statistics of aversive environments. Nat Neurosci. 2015;18(4):590–6.

6. Huang H, Thompson W, Paulus MP. Computational dysfunctions in anxiety: failure to differentiate signal from noise. Biol Psychiatry. 2017;82(6):440–6.

7. Pike AC, Robinson OJ. Reinforcement learning in patients with mood and anxiety disorders vs control individuals: A systematic review and meta-analysis. JAMA psychiatry. 2022;79(4):313–22.

8. Zika O, Wiech K, Reinecke A, Browning M, Schuck NW. Trait anxiety is associated with hidden state inference during aversive reversal learning. Nat Commun. 2023;14(1):4203.

9. Wilson RC, Bonawitz E, Costa VD, Ebitz RB. Balancing exploration and exploitation with information and randomization. Curr Opin Behav Sci. 2021;38:49–56.

10. Schulz E, Gershman SJ. The algorithmic architecture of exploration in the human brain. Current opinion in neurobiology. 2019;55:7–14.

11. Addicott MA, Pearson JM, Wilson J, Platt ML, McClernon FJ. Smoking and the bandit: a preliminary study of smoker and nonsmoker differences in exploratory behavior measured with a multiarmed bandit task. Exp Clin Psychopharmacol. 2013;21(1):66–73.

12. Addicott MA, Pearson JM, Froeliger B, Platt ML, McClernon FJ. Smoking automaticity and tolerance moderate brain activation during explore-exploit behavior. Psychiatry Res. 2014;224(3):254–61.

13. Addicott MA, Pearson JM, Sweitzer MM, Barack DL, Platt ML. A Primer on Foraging and the Explore/Exploit Trade-Off for Psychiatry Research. Neuropsychopharmacology. 2017;42(10):1931–9.

14. Smith R, Taylor S, Wilson RC, Chuning AE, Persich MR, Wang S, et al. Lower levels of directed exploration and reflective thinking are associated with greater anxiety and depression. Frontiers in Psychiatry. 2022;12:782136.

15. Fan H, Gershman SJ, Phelps EA. Trait somatic anxiety is associated with reduced directed exploration and underestimation of uncertainty. Nature Human Behaviour. 2023;7(1):102–13.

16. Lloyd A, McKay RT, Furl N. Individuals with adverse childhood experiences explore less and underweight reward feedback. Proceedings of the National Academy of Sciences. 2022;119(4):e2109373119.

17. Xu Y, Harms MB, Green CS, Wilson RC, Pollak SD. Childhood unpredictability and the development of exploration. Proceedings of the National Academy of Sciences. 2023;120(49):e2303869120.

18. Chorpita BF, Barlow DH. The development of anxiety: The role of control in the early environment. The Neurotic Paradox, Vol 2. 2018:227–64.

19. Nelson CA, Gabard-Durnam LJ. Early adversity and critical periods: neurodevelopmental consequences of violating the expectable environment. Trends in neurosciences. 2020;43(3):133–43.

20. Spadoni AD, Vinograd M, Cuccurazzu B, Torres K, Glynn LM, Davis EP, et al. Contribution of early-life unpredictability to neuropsychiatric symptom patterns in adulthood. Depression and anxiety. 2022;39(10-11):706–17.

21. Biedermann SV, Biedermann DG, Wenzlaff F, Kurjak T, Nouri S, Auer MK, et al. An elevated plus- maze in mixed reality for studying human anxiety-related behavior. BMC Biol. 2017;15(1):125.

22. Walz N, Muhlberger A, Pauli P. A Human Open Field Test Reveals Thigmotaxis Related to Agoraphobic Fear. Biol Psychiatry. 2016;80(5):390–7.

23. Lenow JK, Constantino SM, Daw ND, Phelps EA. Chronic and Acute Stress Promote Overexploitation in Serial Decision Making. J Neurosci. 2017;37(23):5681–9.

24. Teigen K. Yerkes-Dodson: A law for all seasons. Theory and Psychology. 1994;4:525–47.

25. Arnsten AF. Stress weakens prefrontal networks: molecular insults to higher cognition. Nat Neurosci. 2015;18(10):1376–85.

26. Robbins TW, Arnsten AF. The neuropsychopharmacology of fronto-executive function: monoaminergic modulation. Annu Rev Neurosci. 2009;32:267–87.

27. Arnsten A, Robbins T. Neurochemical modulation of prefrontal cortical function in humans and animals. Principles of frontal lobe function2002. Oxford Univeristy Press. 2002: p. 51–84.

28. Arnsten AF. Catecholamine modulation of prefrontal cortical cognitive function. Trends Cogn Sci. 1998;2(11):436–47.

29. Aberg KC, Toren I, Paz R. A neural and behavioral tradeoff underlies exploratory decisions in normative anxiety. Molecular Psychiatry. 2022;27:1573–87.

30. Witte K, Wise T, Huys QJ, Schulz E. Exploring the Unexplored: Worry as a Catalyst for Exploratory Behavior in Anxiety and Depression. PsyArXiv. 2024.

31. Blanco NJ, Otto AR, Maddox WT, Beevers CG, Love BC. The influence of depression symptoms on exploratory decision-making. Cognition. 2013;129(3):563–8.

32. Sheehan DV, Lecrubier Y, Sheehan KH, Amorim P, Janavs J, Weiller E, et al. The Mini-International Neuropsychiatric Interview (MINI): the development and validation of a structured diagnostic psychiatric interview for DSM-IV and ICD-10. Journal of clinical psychiatry. 1998;59(20):22–33.

33. Campbell-Sills L, Norman SB, Craske MG, Sullivan G, Lang AJ, Chavira DA, et al. Validation of a brief measure of anxiety-related severity and impairment: the Overall Anxiety Severity and Impairment Scale (OASIS). Journal of affective disorders. 2009;112(1-3):92–101.

34. Lai HMX, Cleary M, Sitharthan T, Hunt GE. Prevalence of comorbid substance use, anxiety and mood disorders in epidemiological surveys, 1990–2014: A systematic review and meta-analysis. Drug and alcohol dependence. 2015;154:1–13.

35. Bandelow B, Michaelis S. Epidemiology of anxiety disorders in the 21st century. Dialogues in clinical neuroscience. 2015;17(3):327–35.

36. Zhang Z, Mai Y. WebPower: Basic and Advanced Statistical Power Analysis. R package version 0.9.4. 2023. https://CRAN.R-project.org/package=WebPower

37. Starcke K, Wiesen C, Trotzke P, Brand M. Effects of acute laboratory stress on executive functions. Frontiers in Psychology. 2016;7:461.

38. Goldman CM, Takahashi T, Lavalley CA, Li N, Taylor S, Chuning AE, et al. Individuals with Methamphetamine Use Disorder Show Reduced Directed Exploration and Learning Rates Independent of an Aversive Interoceptive State Induction. medRxiv. 2024:2024.05.17.24307491.

39. Spielberger C, Gorsuch R, Lushene R, Vagg P, Jacobs G. Manual for the State-Trait Anxiety Inventory; Palo Alto, CA, Ed. Palo Alto: Spielberger. 1983.

40. Sun Y, Fu Z, Bo Q, Mao Z, Ma X, Wang C. The reliability and validity of PHQ-9 in patients with major depressive disorder in psychiatric hospital. BMC Psychiatry. 2020;20:1–7.

41. Bernstein DP, Stein JA, Newcomb MD, Walker E, Pogge D, Ahluvalia T, et al. Development and validation of a brief screening version of the Childhood Trauma Questionnaire. Child abuse & neglect. 2003;27(2):169–90.

42. Stewart JL, May AC, Poppa T, Davenport PW, Tapert SF, Paulus MP. You are the danger: attenuated insula response in methamphetamine users during aversive interoceptive decision-making. Drug Alcohol Depend. 2014;142:110–9.

43. Kruschwitz JD, Kausch A, Brovkin A, Keshmirian A, Paulus MP, Goschke T, et al. Self-control is linked to interoceptive inference: Craving regulation and the prediction of aversive interoceptive states induced with inspiratory breathing load. Cognition. 2019;193:104028.

44. Berk L, Stewart JL, May AC, Wiers RW, Davenport PW, Paulus MP, et al. Under pressure: adolescent substance users show exaggerated neural processing of aversive interoceptive stimuli. Addiction. 2015;110(12):2025–36.

45. Paulus MP, Flagan T, Simmons AN, Gillis K, Kotturi S, Thom N, et al. Subjecting elite athletes to inspiratory breathing load reveals behavioral and neural signatures of optimal performers in extreme environments. PloS one. 2012;7(1):e29394.

46. Wilson RC, Geana A, White JM, Ludvig EA, Cohen JD. Humans use directed and random exploration to solve the explore–exploit dilemma. Journal of experimental psychology: General. 2014;143(6):2074.

47. Zajkowski WK, Kossut M, Wilson RC. A causal role for right frontopolar cortex in directed, but not random, exploration. Elife. 2017;6:e27430.

48. Waltz JA, Wilson RC, Albrecht MA, Frank MJ, Gold JM. Differential Effects of Psychotic Illness on Directed and Random Exploration. Computational psychiatry (Cambridge, Mass). 2020;4:18–39.

49. Komsta L. outliers: Tests for Outliers. 2022 R package version 0.15, https://CRAN.R-project.org/package=outliers.

50. Hebbali A. olsrr: Tools for Building OLS Regression Models. 2024. R package version 0.6.1.9000, https://olsrr.rsquaredacademy.com, https://github.com/rsquaredacademy/olsrr.

51. Cule E, Moritz S, Frankowski D. ridge: Ridge regression with Automatic Selection of the Penalty Parameter. 2022. R package version 3.3, https://CRAN.R-project.org/package=ridge.

52. Jain AK, Dubes RC. Algorithms for clustering data. Upper Saddle River, NJ: Prentice-Hall, Inc.; 1988.

53. Kuznetsova A, Brockhoff PB, Christensen RHB. lmerTest package: tests in linear mixed effects models. Journal of statistical software. 2017;82:1–26.

54. Kloke JD, McKean JW. Rfit: rank-based estimation for linear models. R J. 2012. Available at: http://rjournal.github.io/archive/2012-2/RJournal_2012-2_Kloke+McKean.pdf.

55. Moser JS, Moran TP, Schroder HS, Donnellan MB, Yeung N. The case for compensatory processes in the relationship between anxiety and error monitoring: a reply to Proudfit, Inzlicht, and Mennin. Front Hum Neurosci. 2014;8:64.

56. Moser JS, Moran TP, Schroder HS, Donnellan MB, Yeung N. On the relationship between anxiety and error monitoring: a meta-analysis and conceptual framework. Front Hum Neurosci. 2013;7:466.

57. Piray P, Daw ND. A model for learning based on the joint estimation of stochasticity and volatility. Nat Commun. 2021;12(1):6587.

58. Heim C, Newport DJ, Mletzko T, Miller AH, Nemeroff CB. The link between childhood trauma and depression: insights from HPA axis studies in humans. Psychoneuroendocrinology. 2008;33(6):693–710.

59. Heim C, Binder EB. Current research trends in early life stress and depression: review of human studies on sensitive periods, gene-environment interactions, and epigenetics. Exp Neurol. 2012;233(1):102–11.

60. Springer KW, Sheridan J, Kuo D, Carnes M. Long-term physical and mental health consequences of childhood physical abuse: results from a large population-based sample of men and women. Child Abuse Negl. 2007;31(5):517–30.

61. Chou K, Wilson RC, Smith R. The influence of anxiety on exploration: A review of computational modeling studies. Neurosci Biobehav Rev. 2024;167:105940.

62. Lachaud L, Jacquet B, Bourlier M, Baratgin J. Mindfulness-based stress reduction is linked with an improved Cognitive Reflection Test score. Frontiers in Psychology. 2023;14:1272324.

63. Calvillo DP, Bratton J, Velazquez V, Smelter TJ, Crum D. Elaborative feedback and instruction improve cognitive reflection but do not transfer to related tasks. Thinking & Reasoning. 2023;29(2):276–304.

64. Orona GA, Trautwein U. Thinking Disposition Education Improves Cognitive Reflection: Experimental Results from An Intervention Study. Thinking Skills and Creativity. 2024;53:101569.

65. Fiorillo D, Papa A, Follette VM. The relationship between child physical abuse and victimization in dating relationships: The role of experiential avoidance. Psychological trauma: theory, research, practice, and policy. 2013;5(6):562.

66. Unger JAM, De Luca RV. The relationship between childhood physical abuse and adult attachment styles. Journal of family violence. 2014;29:223–34.

## References

1. Spielberger C, Gorsuch R, Lushene R, Vagg P, Jacobs G. Manual for the State-Trait Anxiety Inventory; Palo Alto, CA, Ed. Palo Alto: Spielberger. 1983.

2. Norman SB, Hami Cissell S, Means-Christensen AJ, Stein MB. Development and validation of an overall anxiety severity and impairment scale (OASIS). Depression and anxiety. 2006;23(4):245–9.

3. Sun Y, Fu Z, Bo Q, Mao Z, Ma X, Wang C. The reliability and validity of PHQ-9 in patients with major depressive disorder in psychiatric hospital. BMC Psychiatry. 2020;20:1–7.

4. Bernstein DP, Stein JA, Newcomb MD, Walker E, Pogge D, Ahluvalia T, et al. Development and validation of a brief screening version of the Childhood Trauma Questionnaire. Child abuse & neglect. 2003;27(2):169–90.

5. Toplak ME, West RF, Stanovich KE. Assessing miserly information processing: An expansion of the Cognitive Reflection Test. Thinking & reasoning. 2014;20(2):147–68.

6. Bradley MM, Lang PJ. Measuring emotion: the self-assessment manikin and the semantic differential. Journal of behavior therapy and experimental psychiatry. 1994;25(1):49–59.

7. Kalman RE. A new approach to linear filtering and prediction problems. 1960.

8. Waltz JA, Wilson RC, Albrecht MA, Frank MJ, Gold JM. Differential Effects of Psychotic Illness on Directed and Random Exploration. Computational psychiatry (Cambridge, Mass). 2020;4:18–39.

9. Zajkowski WK, Kossut M, Wilson RC. A causal role for right frontopolar cortex in directed, but not random, exploration. eLife. 2017;6.

